# Mutation of SLC7A14 Causes Auditory Neuropathy and Retinitis Pigmentosa Mediated by Lysosomal Dysfunction

**DOI:** 10.1101/2021.06.10.21258486

**Authors:** Kimberlee Giffen, Yi Li, Huizhan Liu, Xiao-Chang Zhao, Chang-Jun Zhang, Ren-Juan Shen, Tianying Wang, Amanda Janesick, Bo-Pei Chen, Shu-Sheng Gong, Bechara Kachar, Zi-Bing Jin, David Z He

## Abstract

Lysosomes contribute to cellular homeostasis via processes including phagocytosis, macromolecule catabolism, secretion, and nutrient sensing mechanisms. Defective proteins related to lysosomal macromolecule catabolism are known to cause a broad range of lysosomal storage diseases. It is unclear, however, if mutations in genes in the autophagy-lysosomal pathway can cause syndromic disease. Here we show that SLC7A14, a transporter protein mediating lysosomal uptake of cationic amino acids, is evolutionarily conserved in vertebrate mechanosensitive hair cells and highly expressed in lysosomes of mammalian cochlear inner hair cells (IHCs) and retinal photoreceptors. Autosomal recessive mutation of *SLC7A14* caused loss of IHCs and photoreceptors, leading to pre-synaptic auditory neuropathy and retinitis pigmentosa in mice and humans. Loss of function mutation altered protein trafficking and disrupted lysosomal homeostasis, resulting in dysregulation of basal autophagy and progressive cell degeneration. This study is the first to implicate autophagy-lysosomal dysfunction in syndromic hearing and vision loss in mice and humans.

## INTRODUCTION

Lysosomes are acidic organelles that serve as catabolic centers in the cell, degrading macromolecules with intraluminal, pH-dependent hydrolases into building blocks to be reused in biosynthesis (1, 2). Lysosomes also function as a signaling hub integrating nutrient-sensing mechanisms and signaling pathways to regulate biogenesis, trafficking, and repair to maintain cellular homeostasis (2, 3). Mutations in genes encoding lysosomal enzymes or transporters leads to accumulation of macromolecules causing multi-system syndromic disorders classified as lysosomal storage diseases (LSDs); to date, over 70 LSDs have been discovered (4). Lysosomal dysfunction can also trigger complex cellular pathogenic cascades, disrupt autophagy, and ultimately lead to cell death. While several LSDs manifest with neurological symptoms, it is unclear if lysosomal dysfunction due to defective genes encoding proteins associated with nutrient sensing mechanisms causes syndromic diseases involving sensory systems such as vision and hearing in humans. Identification of new disease-related genes and their underlying molecular mechanisms is essential for diagnosis, curative therapies and preventative measures.

The mammalian auditory sensory epithelium, the organ of Corti, contains inner and outer hair cells (IHCs and OHCs), each with distinct morphology and function (5). IHCs are the true sensory receptor cells which transmit information to the brain, while the OHCs are a mammalian innovation with the unique capability of changing length in response to changes in receptor potential (6–9). Our previous hair cell type-specific transcriptomic analyses showed that *Slc7a14*, encoding solute carrier family 7 member 14 (SLC7A14), is differentially expressed in IHCs (10–12). SLC7A14 is a cationic amino acid transporter that mediates lysosomal uptake of arginine (13, 14). *Slc7a14* is also expressed in retinal photoreceptor cells and subcortical regions of the brain (13, 15, 16). In 2014, we reported that mutations in *SLC7A14* are responsible for autosomal recessive retinitis pigmentosa (RP) in humans and deletion of *Slc7a14* in adult mice led to decreased retinal thickness and abnormal electroretinography response (15). Knockdown of *slc7a14* in zebrafish showed significant retinal degeneration and visual defects (15).

In this study, we investigated the function of *Slc7a14* in the mammalian inner ear since *Slc7a14* is distinctly expressed in hair cells. We questioned if SLC7A14 is essential for hair cell differentiation, function and survival, and if the point mutation (c.988G>A; p.Gly330Arg) identified in RP patients causes hearing loss. Our study shows that while SLC7A14 was not essential for hair cell differentiation and development, deletion or mutation of *Slc7a14* led to progressive IHC loss in adult mice without affecting OHC function, a condition known as auditory neuropathy. Loss of function mutation also caused thinning of the photoreceptor layer in the mouse retina, a retinopathy indicative of RP. Auditory function tests in patients with *SLC7A14* mutation showed hearing loss in addition to RP. To elucidate the pathogenic mechanisms underlying IHC degeneration, we examined subcellular localization and function of SLC7A14 *in vitro* and *in vivo*. We showed that SLC7A14 was expressed in the lysosomal membrane and trafficked there by a direct intracellular route. The missense mutation in SLC7A14 decreased lysosomal membrane localization and likely disrupted SLC7A14-mediated uptake of arginine into the lysosome. Disruption of lysosomal homeostasis resulted in dysregulation of basal autophagy, causing a significant increase in autophagosome formation and cellular degeneration. Therefore, *SLC7A14* is a novel gene whose mutation can cause hearing and vision impairment as a result of lysosomal dysfunction.

## RESULTS

### Expression of SLC7A14 is distinctive in mature inner hair cells

We first used RNAscope-based smFISH and immunofluorescence to examine the expression of *Slc7a14*/SLC7A14 in developing and adult hair cells of mouse (C57BL/6) cochleae. As shown in Fig. 1a, *Slc7a14* was expressed in both IHCs and OHCs at postnatal day 0 (P0). However, at P7 there was a distinctive downregulation in OHCs, while expression continued to increase in IHCs (Fig. 1a, b). IHC-specific expression of SLC7A14 increased from P12, and strong expression was observed in P30 and P60 IHCs extending from the cochlear base to apex (Fig. 1b, c; Supplementary Fig. 1a). Expression of *Slc7a14* mRNA was observed in the spiral ganglion at P0; however, no protein expression was detected at any age (Supplementary Fig. 1b, c). Consistent with previous observations, expression of SLC7A14 increased in the developing mouse retina, and was restricted to rod and cone photoreceptors and ganglion neurons in the adult retina (15) (Supplementary Fig. 1d). In addition to expression in specialized sensory receptors in the cochlea and retina, SLC7A14 is expressed in distinct neuronal populations in the brain including the hippocampus (Supplementary Fig. 1e-f). Mild gene level expression was observed in vestibular hair cells at embryonic day 18.5; however, no mRNA or protein expression was detected in postnatal vestibular hair cells (Supplementary Fig. 2).

**Fig. 1:**
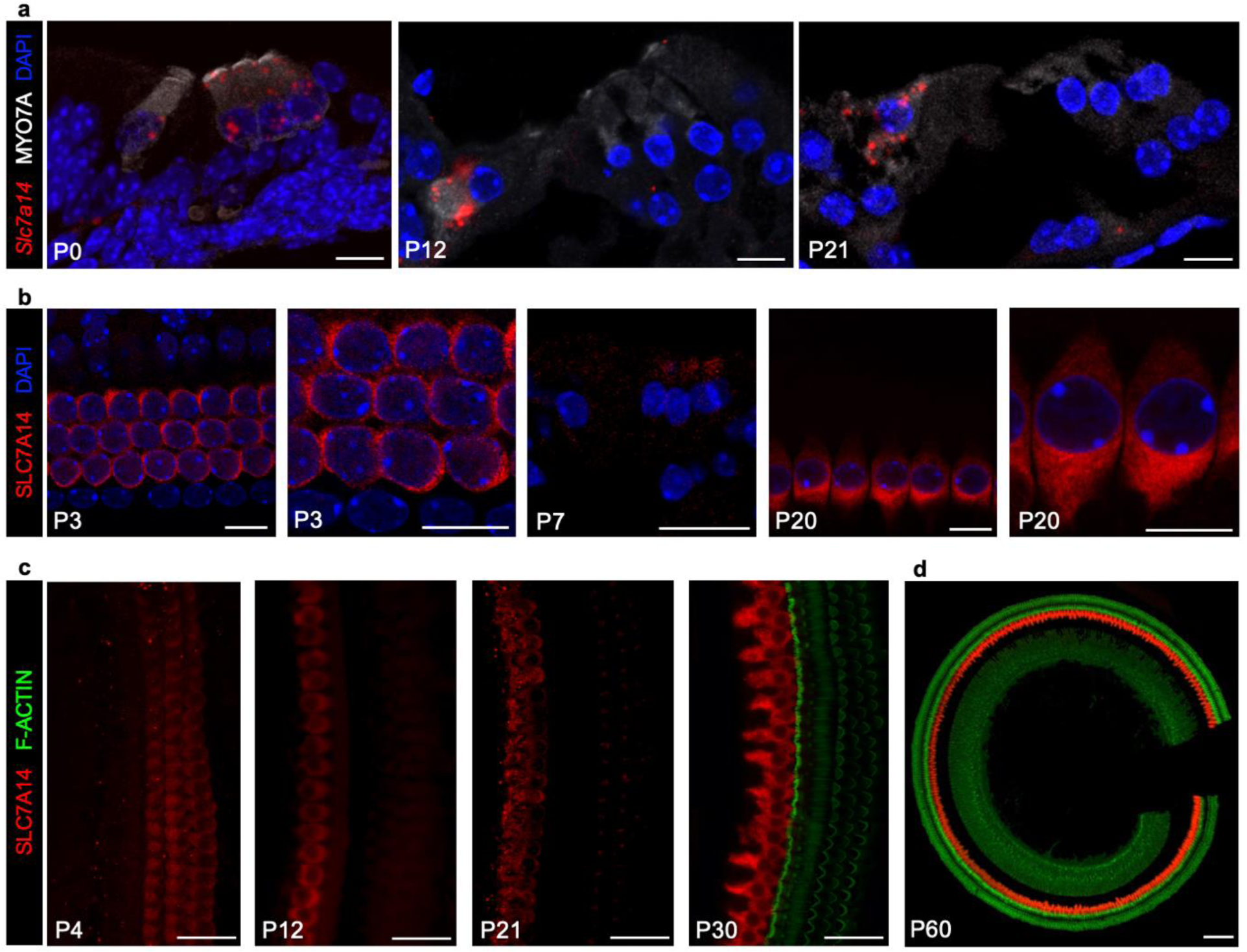
Expression of *Slc7a14*/SLC7A14 in developing mouse organ of Corti. (a) RNAscope smFISH of *Slc7a14* (red probe) in developing mouse organ of Corti, with IHCs and OHCs identified based on MYO7A expression (Bars: 10 μm). (b) SLC7A14 expression in mouse (C57BL/6) OHCs observed at P3 and P7, and distinctive expression in IHCs at P20 (Bars: 10 μm). (c) Immunofluorescent labelling of SLC7A14 in wildtype IHCs and OHCs from P4 to P30 (Bars: 20 μm). (d) Expression of SLC7A14 observed in mature P60 IHCs (Bar: 100 μm).

Immunofluorescence was then used to examine cell-type specific expression of SLC7A14 orthologs in mature vertebrate auditory end organs to demonstrate conservation of this relatively unknown SLC protein. Slc7a14(b) expression was observed in zebrafish inner ear hair cells in the saccule and utricle, as well as the lateral line neuromasts, consistent with our previous RNA-seq data (17) (Fig. 2a, b). Similarly, hair cells in the turtle auditory papilla and chicken basilar papilla exhibited high-level expression of Slc7a14 (Fig. 2c, d). Expression of SLC7A14 in the mammalian cochlea was observed to be IHC-specific in mouse and rat, and most importantly, in the human cochlea (Fig. 2e-g). Thus, SLC7A14 is highly conserved through evolution, which is evidenced by homologous protein expression in the sensory hair cells across vertebrate species and more specifically in mammalian IHCs.

**Fig. 2:**
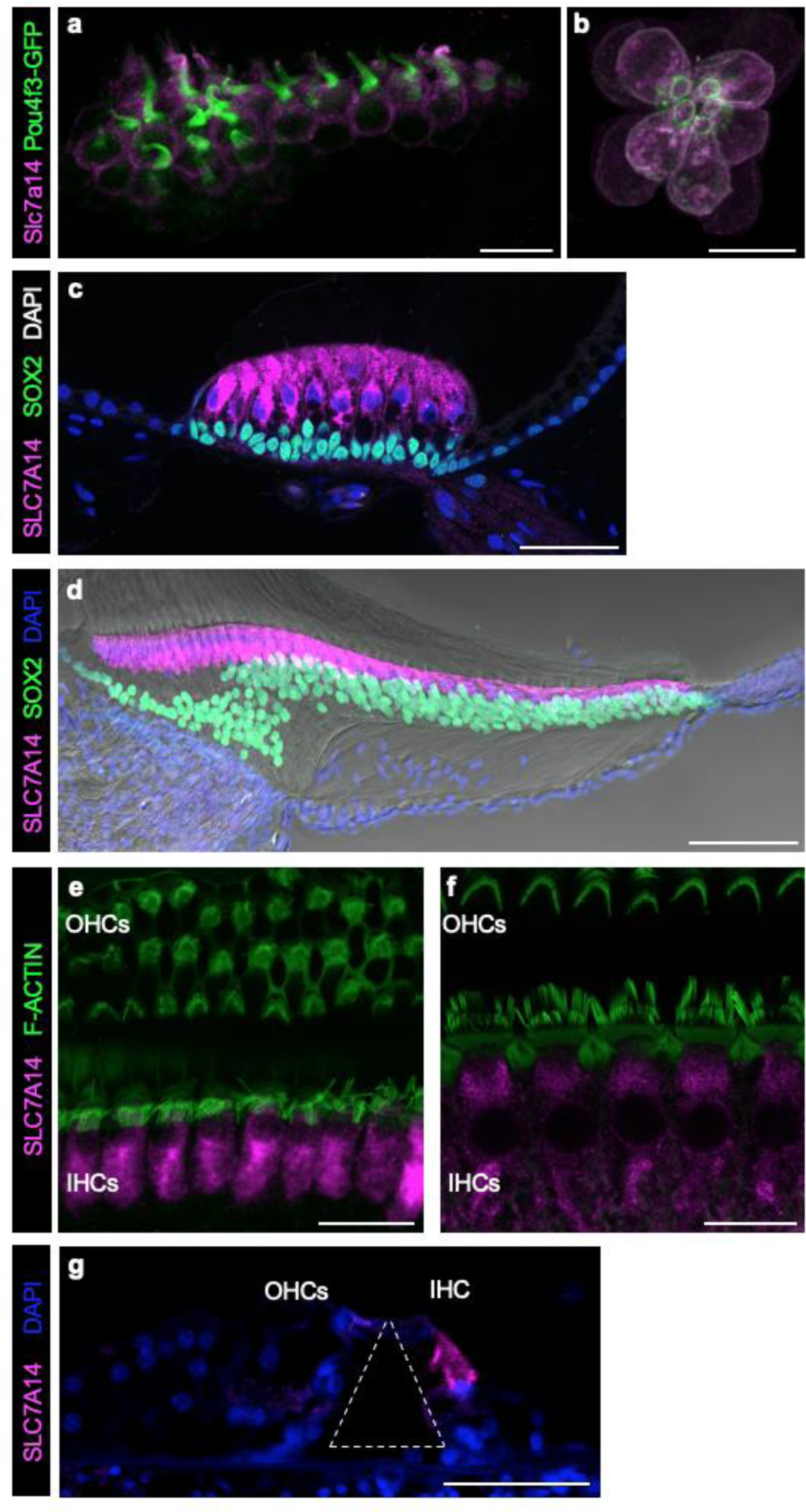
Conserved expression of SLC7A14 orthologs among vertebrate sensory hair cells. SLC7A14 labeled by immunofluorescence in (a) zebrafish saccule and (b) neuromast hair cells (72 hours past fertilization; Bars: 10 µm); (c) turtle auditory papilla hair cells (Adult; Bar: 20 µm); (d) chicken basilar papilla hair cells (7 days post-hatch; Bar: 50 µm); (e) mouse cochlear hair cells (P60; Bar: 20 µm); (f) rat cochlear hair cells (P20; Bar: 20 µm), and adult human cochlear IHC (tunnel of Corti identified by dashed white line) (Bar: 50 µm).

### Loss of SLC7A14 function leads to progressive hearing loss in adult mice

The progressive loss of photoreceptors and visual dysfunction, indicative of RP, was previously observed in *Slc7a14* knockout (KO) mice (15). We sought to determine if loss of SLC7A14 function would also lead to hearing loss in *Slc7a14* KO mice, which lack SLC7A14 expression in the IHCs (Supplementary Fig. 3). Auditory function in mice was characterized by measuring thresholds of auditory brainstem response (ABR) and distortion product otoacoustic emission (DPOAE). ABRs are electrical signals evoked from the brainstem during presentation of an acoustic signal, while DPOAEs are generated by the cochlear amplifier and reflect OHC function (6, 7). Fig. 3 shows the ABR thresholds of homozygous KO (*Slc7a14*^-/-^), heterozygous (*Slc7a14*^+/-^) and wildtype (*Slc7a14*^+/+^) mice at 1, 2, 3 and 6 months of age. At one month, ABR thresholds of KO mice were comparable to wildtype and heterozygous mice, suggesting that mechanotransduction, amplification and synaptic transmission of hair cells are normal at this stage (Fig. 3a). Thus, deletion of *Slc7a14* did not affect differentiation and development of IHCs or OHCs. At 3 months the ABR thresholds of KO mice were significantly elevated at all frequencies tested. Thresholds were further elevated at 6 months, suggesting progressive hearing loss in the KO mice (Fig. 3a, b). Notably, the ABR thresholds of heterozygous mice remained comparable to wildtype mice at all ages indicating that a single functional allele provides sufficient protein for IHC function. We also measured DPOAE in these mice to assess the motor activity of OHCs. Contrary to the ABR results, there was no measurable difference in DPOAE thresholds in KO, heterozygous and wildtype mice at 3 and 6 months (Fig. 3c), suggesting that OHCs are spared despite deletion of this gene.

**Fig. 3:**
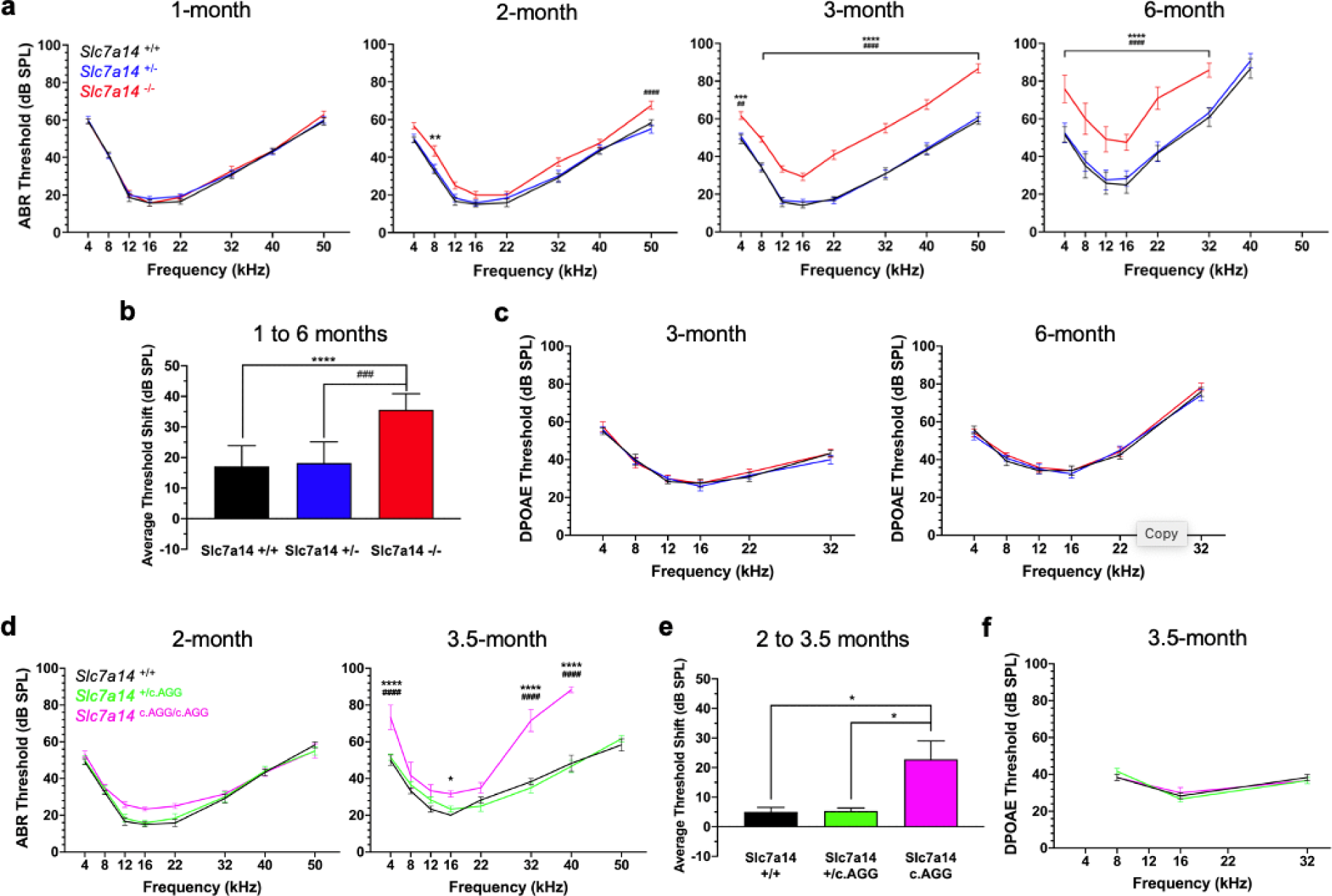
Measurement of auditory function in *Slc7a14* knockout and knockin mice. (a) ABR threshold measurements of *Slc7a14*^+/+^ (black line), *Slc7a14*^+/-^ (blue line), and *Slc7a14*^-/-^ (knockout, red line) mice at 1, 2, 3, and 6 months of age (n = 6). (b) ABR threshold shifts at 6 months compared to 1-month baseline. (c) DPOAE threshold measurements of *Slc7a14* ^+/+^, *Slc7a14* ^+/-^, and *Slc7a14* ^-/-^ mice at 3 and 6 months (n = 6). (d) ABR thresholds of *Slc7a14* ^+/+^ (black line), *Slc7a14* ^+/c.AGG^ (green), and *Slc7a14* ^c.AGG/c.AGG^ (knockin, magenta line) mice at 2 month (n = 3) and 3.5 months of age (n = 6). (e) ABR threshold shifts in knockin mice at 3.5-months compared to 2- month baseline. (f) DPOAE threshold measurements in *Slc7a14* ^+/+^, *Slc7a14* ^+/ c.AGG^, and *Slc7a14* ^c.AGG/c.AGG^ mice at 3.5 months (n WT = 6, KI = 3). Symbols: Wildtype to KO/ KI * p < .05, ** p < .005, *** p < .0005, **** p < .0001; Heterozygous to KO/ KI ## p < .005, ### p < .0005, #### p < .0001; and, error bars: ±SE.

Previously, we reported that *SLC7A14* mutations were linked to autosomal recessive RP, including the missense variant c.988G>A (15). To demonstrate that this mutation would lead to hearing loss in mice, we used CRISPR/Cas9 nuclease to generate *Slc7a14* knockin (KI) mice with the missense codon substitution (c.AGG) encoding SLC7A14 p.Gly330Arg (Supplementary Fig. 3). Immunostaining with anti-SLC7A14 antibody showed positive, albeit weaker expression of SLC7A14 in the IHCs of homozygous KI mice (Supplementary Fig. 3). Auditory function was examined in wildtype, heterozygous (*Slc7a14*^+/c.AGG^) and KI (*Slc7a14*^c.AGG/c.AGG^) mice at 2 and 3.5 months of age. There was no significant difference in ABR thresholds among the three genotypes at 2 months (Fig. 3d). At 3.5 months, ABR threshold was significantly elevated at 4, 32, and 40 kHz in KI mice compared to wildtype and heterozygous mice (Fig. 3d, e). Similar to heterozygous KO mice, the auditory threshold of heterozygous KI mice remained comparable to that of the wildtype mice. Additionally, DPOAE thresholds at 3.5 months showed no difference among the three genotypes, consistent with that observed in the KO mice (Fig. 3f).

### Degeneration of IHCs was observed in *Slc7a14* KO and KI mice

Scanning electron microscopy (SEM) was used to examine hair bundle morphology in KO and KI mice. Fig. 4 shows sample images of IHC and OHC stereocilia bundles in the mid-cochlear region of KO (3-month-old) and KI (3.5-month-old) mouse cochleae. As shown, the stereocilia of IHCs exhibited signs of absorption, fusion, recession in both KO and KI mice. In addition to IHC degeneration, sporadic loss of IHC stereocilia bundles was also observed (Fig. 4b, c marked by yellow arrows). There was no region-specific pattern of IHC degeneration as signs of sporadic IHC stereocilia bundle degeneration and loss were observed along the length of the cochlea. Not surprisingly, OHC bundles of the KO and KI mice appeared to be normal with a V-shaped appearance and no clear signs of degeneration and loss (Fig. 4).

**Fig. 4:**
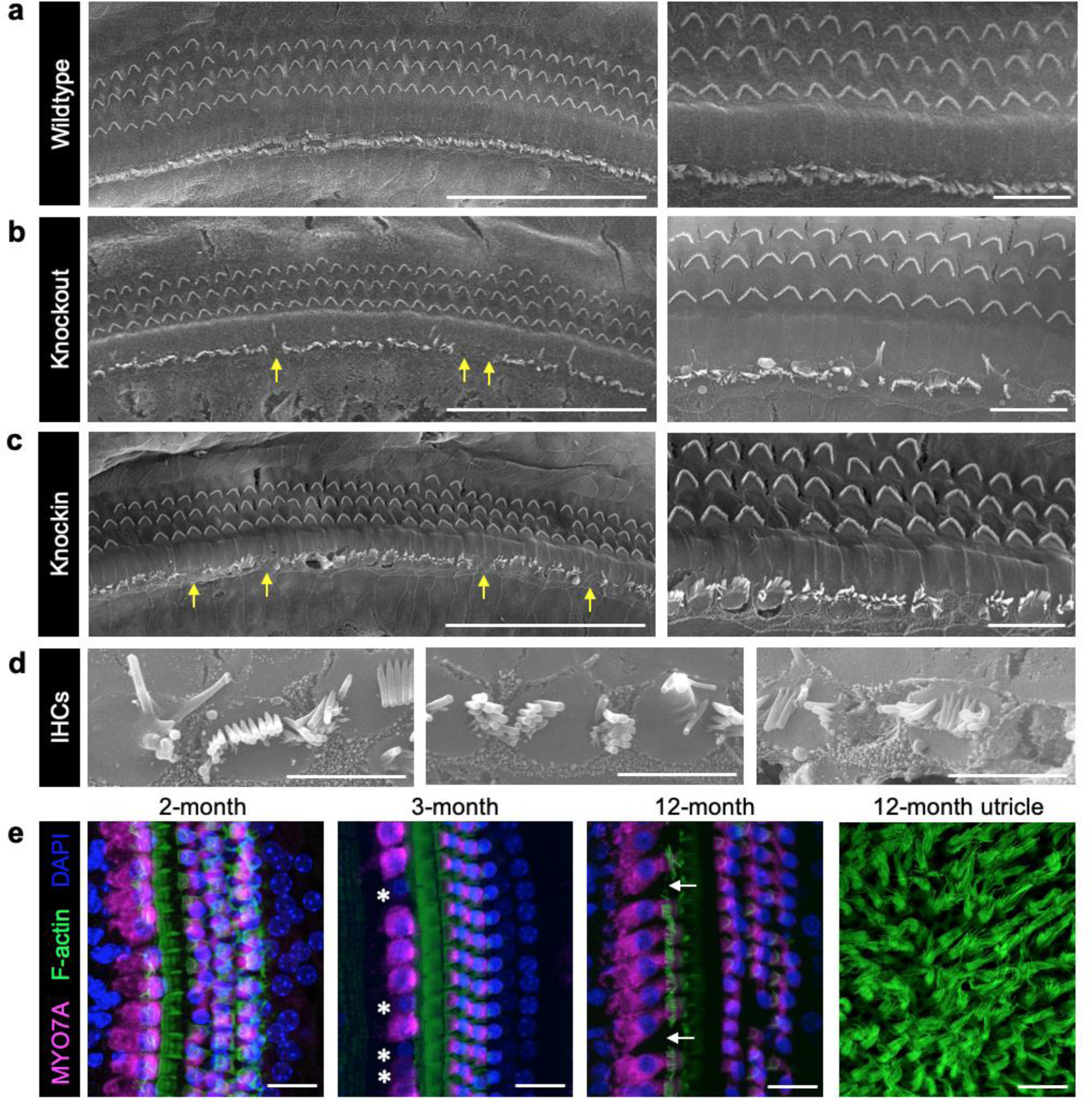
Morphology of knockout and knockin mouse cochlear hair cells. Scanning electron microscopy of 3-month old (a) wildtype cochlea, (b) *Slc7a14*^-/-^ (knockout) mouse cochlea, and (c) *Slc7a14^c.AGG/c.AGG^* (knockin) mouse cochlea (Bars: 50 μm, 10 μm). Yellow arrows indicate absent IHC bundles. (d) Representative images of IHC stereocilia bundles from middle turn of knockout mouse cochlea show clear signs of degeneration, fusion, and absorption (Bars: 5 μm). (e) Immunofluorescence of knockout mouse cochlea at 2, 3, and 12 months. Asterisk indicates decreased MYO7A expression; white arrow indicates absent IHCs. Right panel: F-actin labeled utricle hair cell stereocilia bundles in 12-month knockout mouse (Bars: 20 μm).

Confocal microscopy was used to examine hair cell survival in KO mice. Immunostaining of KO mouse cochlea showed decreased MYO7A expression (sign of degeneration) in IHCs at 3 months, while the middle turn of 12-month-old KO cochlea had absent IHCs (Fig. 4e). Consistent with lack of SLC7A14 expression in vestibular hair cells, stereocilia bundles of utricle hair cells appeared normal and no missing hair cells were observed in a 12-month-old KO mouse (Fig. 4e). We also compared the number of synaptic ribbons in 3.5-month-old KI and wildtype IHCs using anti-CtBP2 antibodies. CtBP2 is a major component of synaptic ribbons and immunostaining against CtBP2 is often used to quantify presynaptic ribbons. We found no measurable difference between the number of CtBP2-positive puncta per IHC in the apical turn (KI = 8.61 ± 1.70; WT = 8.46 ± 2.32; p = 0.83) or middle turn (KI = 15.7 ± 0.87; WT = 14.2 ± 1.19; p = 0.21).

### The SLC7A14 point mutation causes syndromic disease

To confirm the syndromic phenotype associated with mutation of *Slc7a14*, we conducted histochemical analysis of cross-sections of KI mouse retinas. The KI retina was thinner compared to the age matched wildtype retina, with a visible difference in the thickness of the photoreceptor layer (Fig. 5a, b). As a complimentary approach, we also examined whether *SLC7A14* mutations cause hearing loss in humans by measuring auditory function in three proband patients diagnosed with autosomal recessive RP due to the homozygous mutation *SLC7A14* (c.988G>A) (15). The patients live in rural communities and did not have any reported history of job-related exposure to noise. After tympanometry showed normal middle ear function, each patient was tested with pure-tone audiometry (Fig. 5c-e). Patient 1, a male aged between 36 and 41, displayed moderate to severe bilateral hearing loss at 4 and 8 kHz. Patient 2, a male aged between 5 and 10, had mild bilateral hearing loss between 1 and 8 kHz. The third patient, a female aged between 45 and 50, showed normal hearing between 1 and 8 kHz; however, extended-frequency testing showed moderate to severe bilateral hearing loss between 9 and 13 kHz. Additionally, all three patients were subjected to DPOAE measurements between 1 and 8 kHz to assess OHC function. The otoacoustic emissions measured were diagnosed as present and normal, an example is presented in Fig. 5f. Retina morphology and function of the patients were examined. Fundoscopy showed bone-spicule pigment deposits in the peripheral retina in the representative images from two patients (Fig. 5g, h). Optical coherence tomography (OCT) showed marked disruption of the photoreceptor layer of the retina with preservation of the ellipsoid zone in the fovea in patient 1; OCT scans of patient 2 revealed loss of photoreceptor layer outside the macula, and bilateral macular edema (Fig. 5g, h). Finally, electroretinography (ERG) showed severe decreased scotopic and photopic wave amplitude (Fig. 5i). These examinations suggest retinal degeneration of both eyes in these patients. Collectively, the auditory and visual phenotypes characterized in the patients harboring the loss of function variant *SLC7A14* (c.988G>A) confirms syndromic disease.

**Figure 5:**
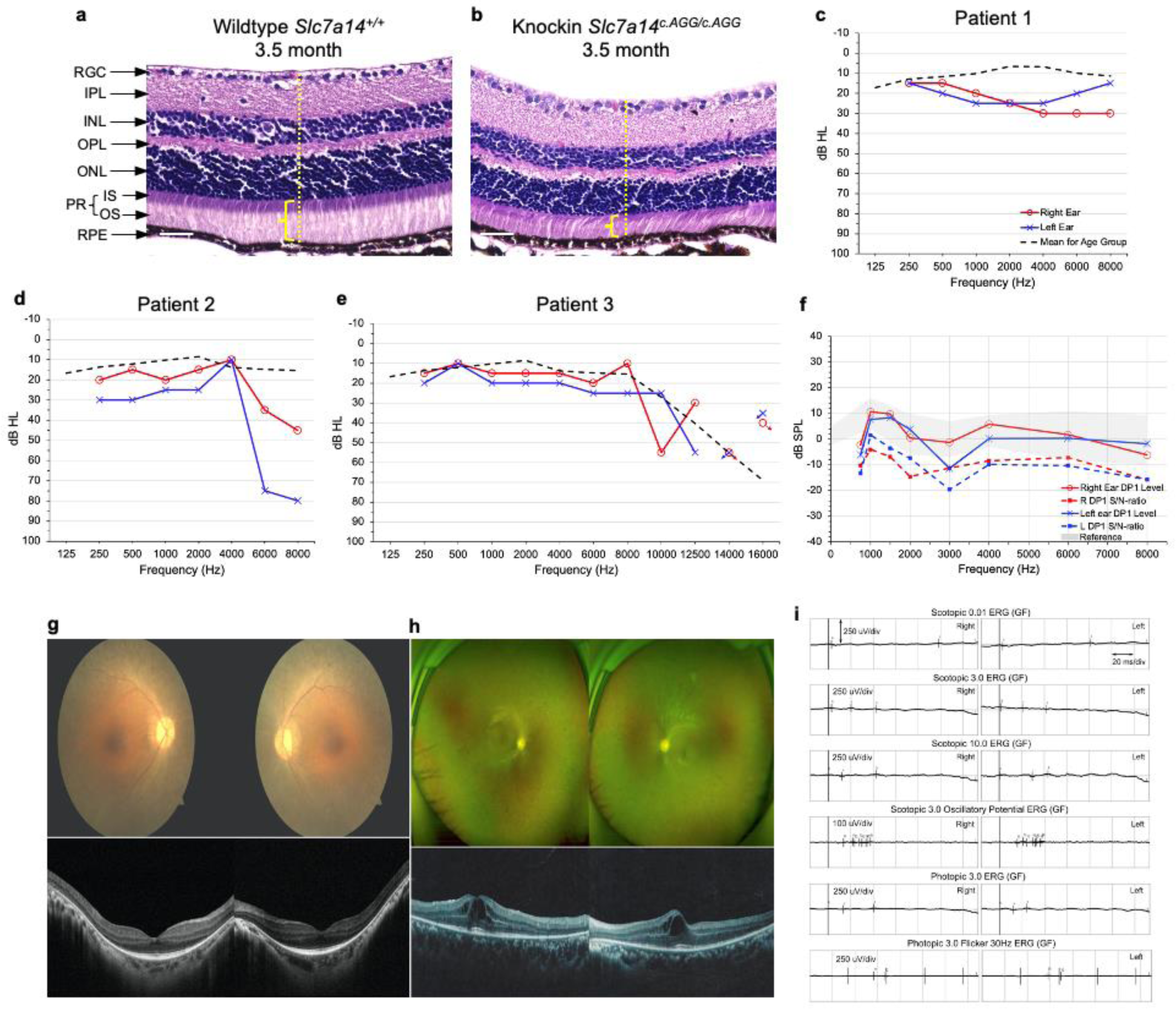
Syndromic disease associated with *SLC7A14* loss of function mutation. HE stained retinal cross sections from 3.5-month-old (a) wildtype and (b) homozygous knockin mice. Decreased retinal thickness (yellow dotted line) and thinning of the photoreceptor layer (yellow bracket) was observed in knockin mouse retina. Retinal layers: RGC: Retinal ganglion cells; IPL: inner plexiform layer; INL: inner nuclear layer; OPL: outer plexiform layer; ONL: outer nuclear layer; PR: photoreceptor layer, IS: inner segment and OS: outer segment of rods and cones; RPE: retinal pigment epithelium. Bars: 50 μm. (c-e) Audiogram measurements of three probands with the *SLC7A14* c.988G<A mutation. Threshold levels are shown in decibels in hearing level (dB HL) for the left and right ears, with the age matched mean dB HL shown for comparison. Disconnected data points with arrow indicates no response. (f) Representative DPOAE measurement. F1 level: 65 dB; F2 level: 55 dB. Noise floor indicated as Signal/Noise ratio, shown as dotted line. Normal range shaded in gray. (g) Fundoscopic images and OCT scans of the retinas of Patient 1. (h) Ultra-wide field fundus images and OCT scans of Patient 2. (i) Full-field EGR of the two patients.

### SLC7A14 is trafficked by direct intracellular routes to the lysosomal membrane

While SLC7A14 is highly and specifically expressed in IHCs, the transporter’s subcellular localization and function in sensory IHCs is unknown. Previous *in vitro* studies showed that SLC7A14 localizes to the membranes of intracellular vesicles, including lysosomes (14, 15). We used two approaches to examine subcellular location of SLC7A14 expression. We first used anti- SLC7A14 antibodies and co-labelled different intracellular organelles (i.e., endoplasmic reticulum with anti-calreticulin, Golgi apparatus with anti-GLG1, and lysosomes with anti-LAMP1 or anti- LAMP2) to examine trafficking and subcellular localization of SLC7A14 in SH-SY5Y neuroblastoma cells, which have moderate endogenous expression of the protein. Colocalization analysis did not include mitochondria as SLC7A14 was not predicted nor previously shown to localize to the mitochondrial membrane (14, 15). Confocal microscopy and colocalization analyses showed SLC7A14 expression co-localized with the expression of LAMP1 or LAMP 2 (Fig. 6c, d), suggesting its localization to lysosomes; however, there was also labeling of SLC7A14 that did not coincide with the lysosome. SLC7A14 exhibited peri-nuclear expression with Calreticulin and GLG1 indicating that the protein also localizes to the endoplasmic reticulum and Golgi (Fig. 6a, b). Nikon NIS-Elements software was used to measure the Pearson’s correlation coefficient (PCC), which quantifies the overlap of each respective fluorophore-labelled protein (e.g., LAMP1 or Calreticulin) and SLC7A14 (18). The mean PCC values ranged from 0.61 to 0.71, with the highest overlap between lysosomes expressing SLC7A14 and LAMP1 (Fig. 6e). The lower, albeit still significant, correlation between SLC7A14 and GLG1 suggests that the presence of the protein in the Golgi is more transient, whereas SLC7A14 is more abundant in the endoplasmic reticulum, where it is produced, and at its final destination, the lysosome. Additionally, SLC7A14 was not expressed on the plasma membrane suggesting that the membrane bound transporter is trafficked by a direct intracellular route to the lysosome (19).

**Fig. 6:**
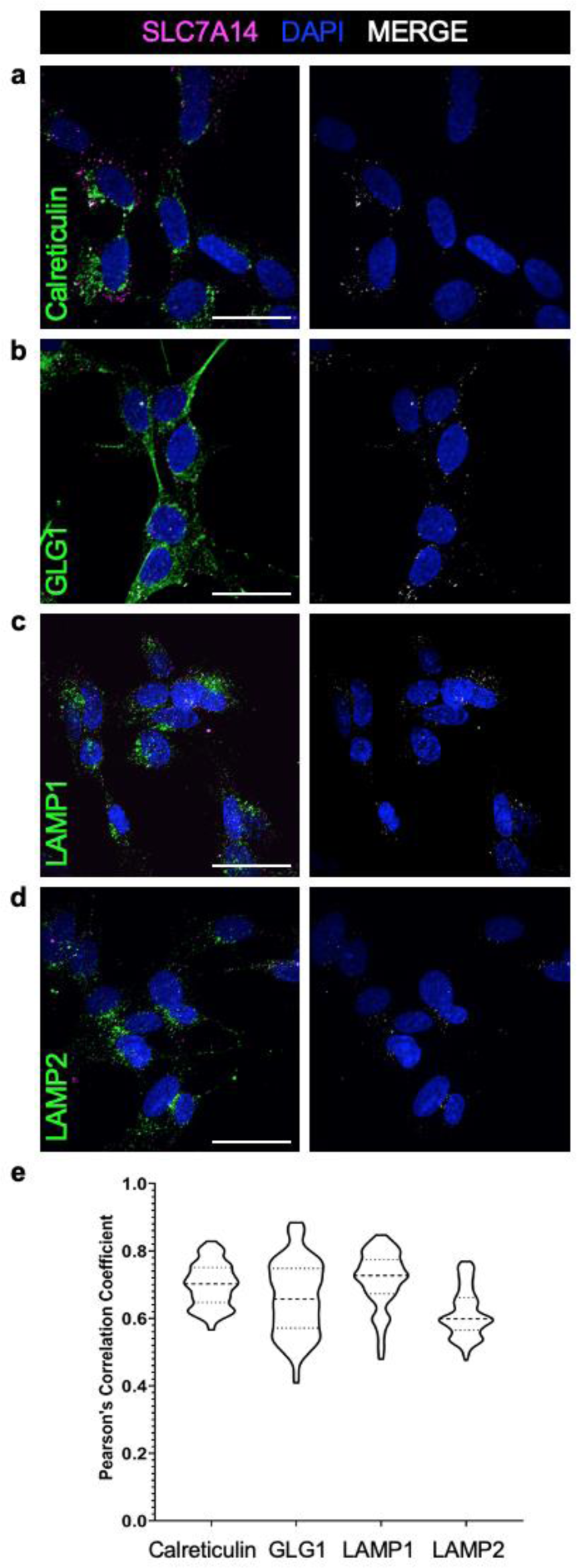
Colocalization of SLC7A14 within intracellular membranes. Immunofluorescence of SH-SY5Y cells shows colocalization of endogenous SLC7A14 and (a) Calreticulin to label the endoplasmic reticulum; (b) GLG1 to label the golgi body; and, (c) LAMP1 and (d) LAMP2 to label the lysosomes (Bars: 10 μm). (e) Violin plot of colocalization of each respective organelle membrane label and SLC7A14 quantified by Pearson’s correlation coefficient. Mean of individual PCC values for > 100 z-stack images (n = 3 technical replicates per label) were compared by one-way ANOVA. P values were calculated by post-hoc analysis with the Tukey test; (*α* = .05); (**** p < .00001).

### Mutant SLC7A14 protein showed aberrant subcellular localization

Membrane targeting is essential for the function of the membrane bound SLC protein. We questioned whether the pathogenicity of the missense mutation SLC7A14 p.Gly330Arg, which alters the primary protein structure, could result in the reduction or loss of lysosomal membrane expression. To demonstrate that mutant SLC7A14 shows aberrant cellular localization, SH-SH5Y cells were transfected with pcDNA3.1 plasmids encoding either the human wildtype or mutant SLC7A14 p.Gly330Arg protein with a fused eGFP tag at the C-terminus. Following transfection, live cells were labeled with LysoTracker Red and observed for localization of SLC7A14-eGFP to lysosomes (Fig. 7a, b). The eGFP tag did not alter normal trafficking of the wildtype protein to the lysosome. However, there was a significant decrease in localization of the SLC7A14 p.Gly330Arg -eGFP protein to the lysosome (Fig. 7c). Next, subcellular localization of the mutant SLC7A14 p.Gly330Arg protein was examined in the KI mouse cochleae. Similarly, immunostaining showed that colocalization of SLC7A14 p.Gly330Arg with each respective organelle label was significantly decreased in the KI IHCs compared to the wildtype IHCs (Fig. 8). The aberrant localization and diffuse cytosolic expression suggested that the modified residue in the loop domain between transmembrane domains 7 and 8 may disrupt trafficking to the lysosome, though it is unclear how the glycine to arginine substitution interrupts protein folding, stability, and membrane targeting. Furthermore, the abnormal localization of the lysosomal transporter resulting from the missense mutation provides some explanation for the functional deficit seen in KI mice and autosomal recessive RP patients.

**Fig. 7:**
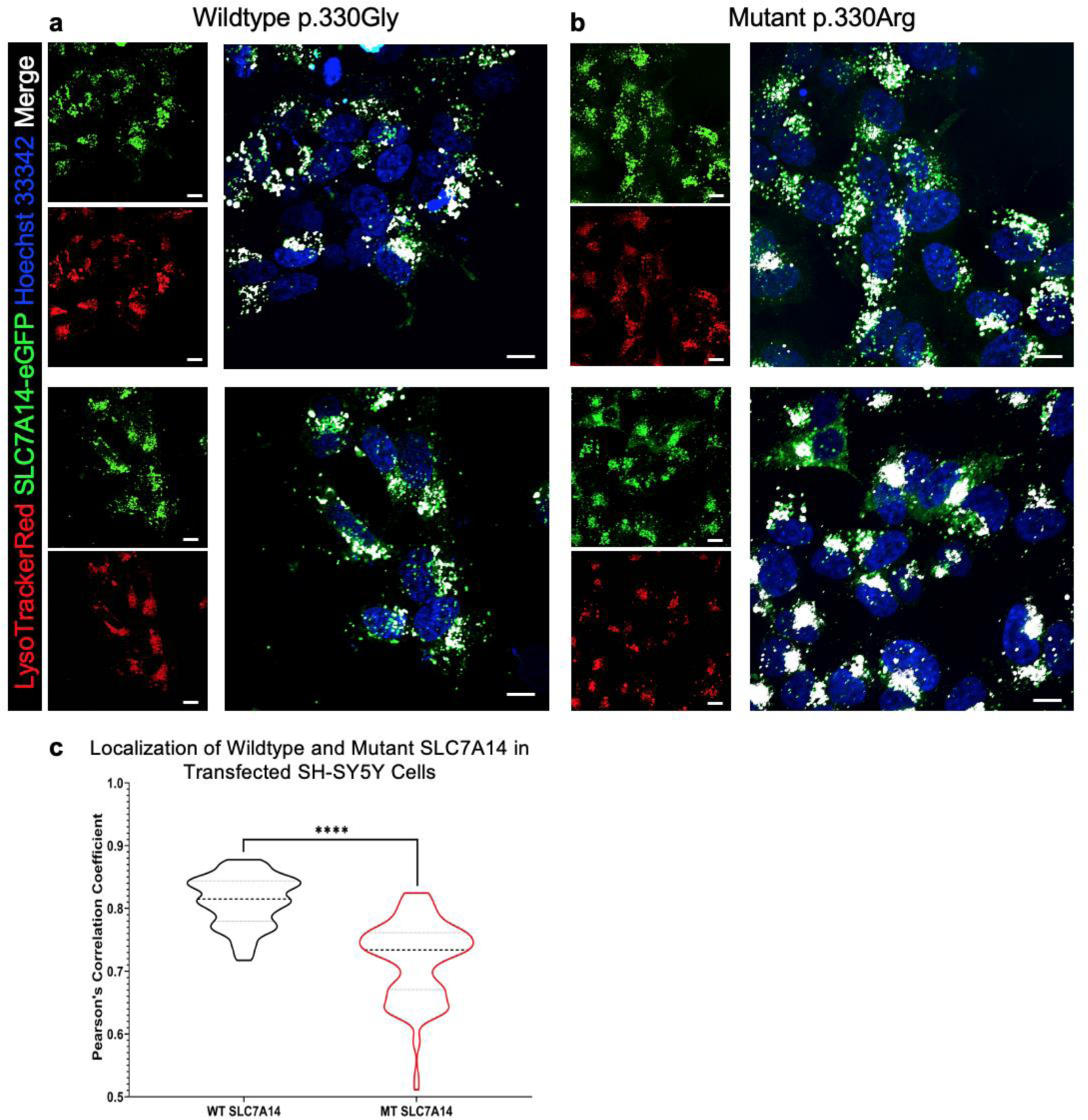
Localization of wildtype and mutant SLC7A14 in SH-SY5Y cells. Live SH-SY5Y cells were transfected with (a) wildtype or (b) mutant p.330Gly>Arg SLC7A14 plasmids and then LysotrackerRed99 and Hoechst 33342 labelled lysosomes and nuclei, respectively (Bars: 10 μm). Colocalization between SLC7A14-eGFP and Lysotracker shown in white. (c) Violin plots of measured overlap of green and red channels based on Pearson’s correlation coefficient. Mean of individual PCC values for > 125 z-stack images (n = 3 WT or MT transfected technical replicates) P value calculated with unpaired, two-tailed t-test; (*α* = .05); (**** p < .00001).

**Fig. 8:**
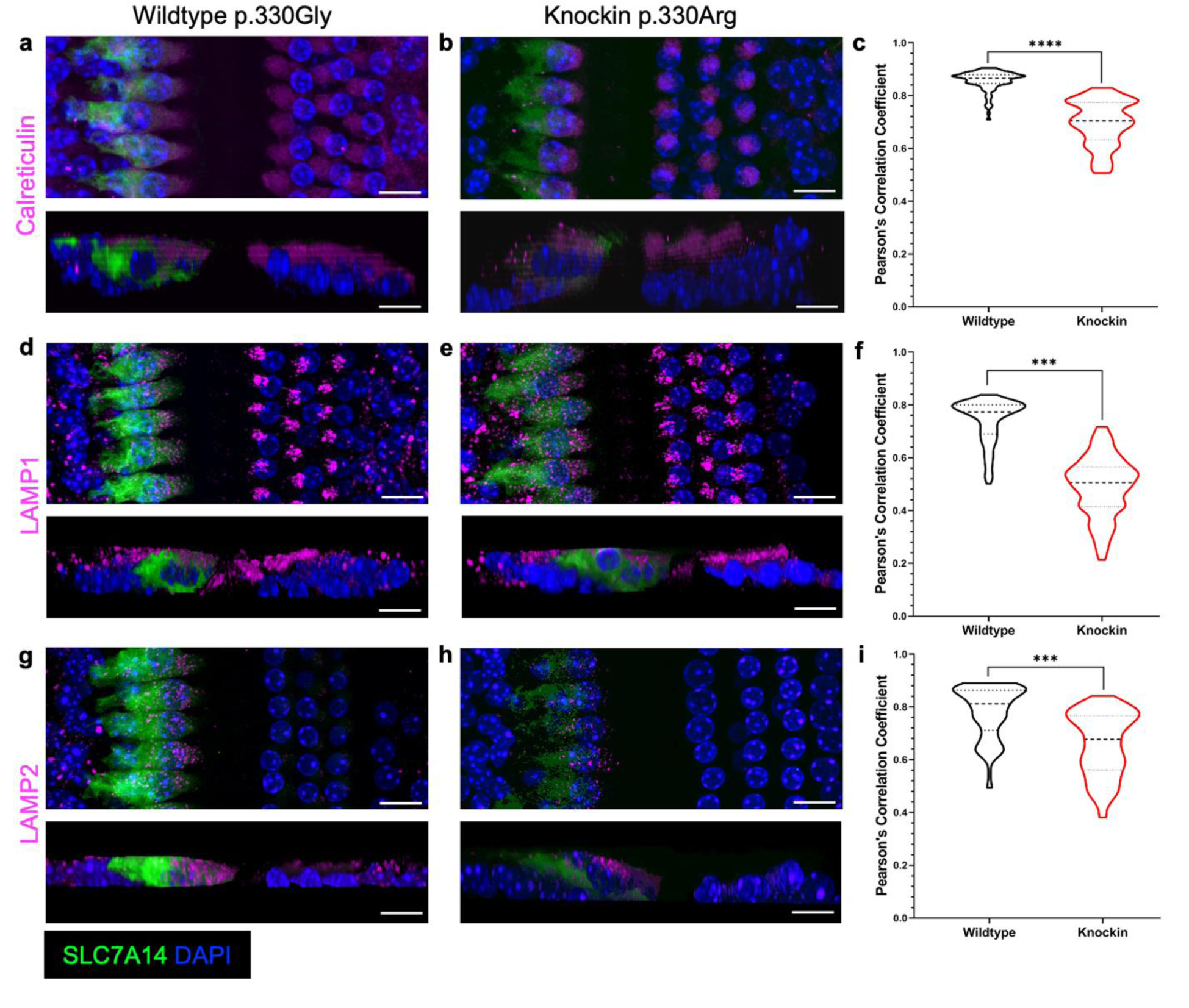
Localization of SLC7A14 in wildtype and knockin inner hair cells. Immunofluorescent labelling of SLC7A14 and respective organelle labels: (a, b) endoplasmic reticulum marker Calreticulin, (d, e) lysosome label LAMP1, and (g, h) lysosome label LAMP2. (Bars: 10 μm) (c, f, and i) Violin plots of Pearson’s correlation coefficient used to quantify colocalization of each organelle label in wildtype and knockin IHCs for > 85 z-stack images (n = 3 technical replicates per genotype). P values for comparison of mean PCC values between wildtype and knockin mouse IHCs were obtained by one-way ANOVA, with post-hoc analysis for multiple comparisons using the Sidak test (*α* = .05); (*** p < 0.0001; **** p < .00001).

### The SLC7A14 mutation results in dysregulation of basal autophagy

Lysosomal SLC transporters have frequently been implicated in disease; their dysfunction can result in impairment of autophagy or lysosomal storage disorders (20, 21). The activity of SLC7A14 in the lysosomal uptake of arginine suggests that the transporter may regulate metabolic homeostasis and autophagy (14, 22). In post-mitotic cells such as photoreceptors, hair cells, and neurons, basal autophagy is an essential mechanism for cell survival, and dysregulation of autophagy in these cell populations can lead to cell degeneration and death (23). For this reason, we first investigated the role of SLC7A14 in regulating autophagy by examining autophagosome formation and autophagy activity in SH-SY5Y cells after siRNA-mediated knockdown of *SLC7A14.* Cultured SH-SY5Y cells were transfected with *SLC7A14* siRNA, control siRNA, or incubated in transfection media alone. Following gene knockdown, cells were incubated in normal serum or autophagy-inducing starvation conditions. Quantitative PCR and Western blot were used to measure mRNA and protein expression, respectively. The *SLC7A14* siRNA treated cells had decreased mRNA and protein expression compared to untreated and control siRNA treated SH-SY5Y cells (Fig. 9a-c). There was a significant increase in the LC3-II to LC3-I ratio in both untreated and control siRNA-treated cells after starvation, indicative of increased autophagosome formation and autophagy activity (Fig. 9d, e). However, compared to the untreated cells, the SH-SY5Y cells treated with *SLC7A14* siRNA had a significant increase in LC3-II to LC3-I ratio under normal serum conditions rather than following starvation. These results suggest that decreased expression of SLC7A14 increases basal autophagy. To confirm these findings *in vivo*, basal autophagy activity was examined in KI and KO mouse cochleae (aged 1.5 to 3 months). The number of LC3 puncta in IHCs, marking active autolysosome formation, was significantly higher in KI and KO mice, compared to wildtype animals (Fig. 9f, g). Dysfunction of autophagy in mature hair cells, due to loss of normal SLC7A14 function, suggests a pathological mechanism leading to progressive IHC degeneration and subsequent hearing loss.

**Fig. 9:**
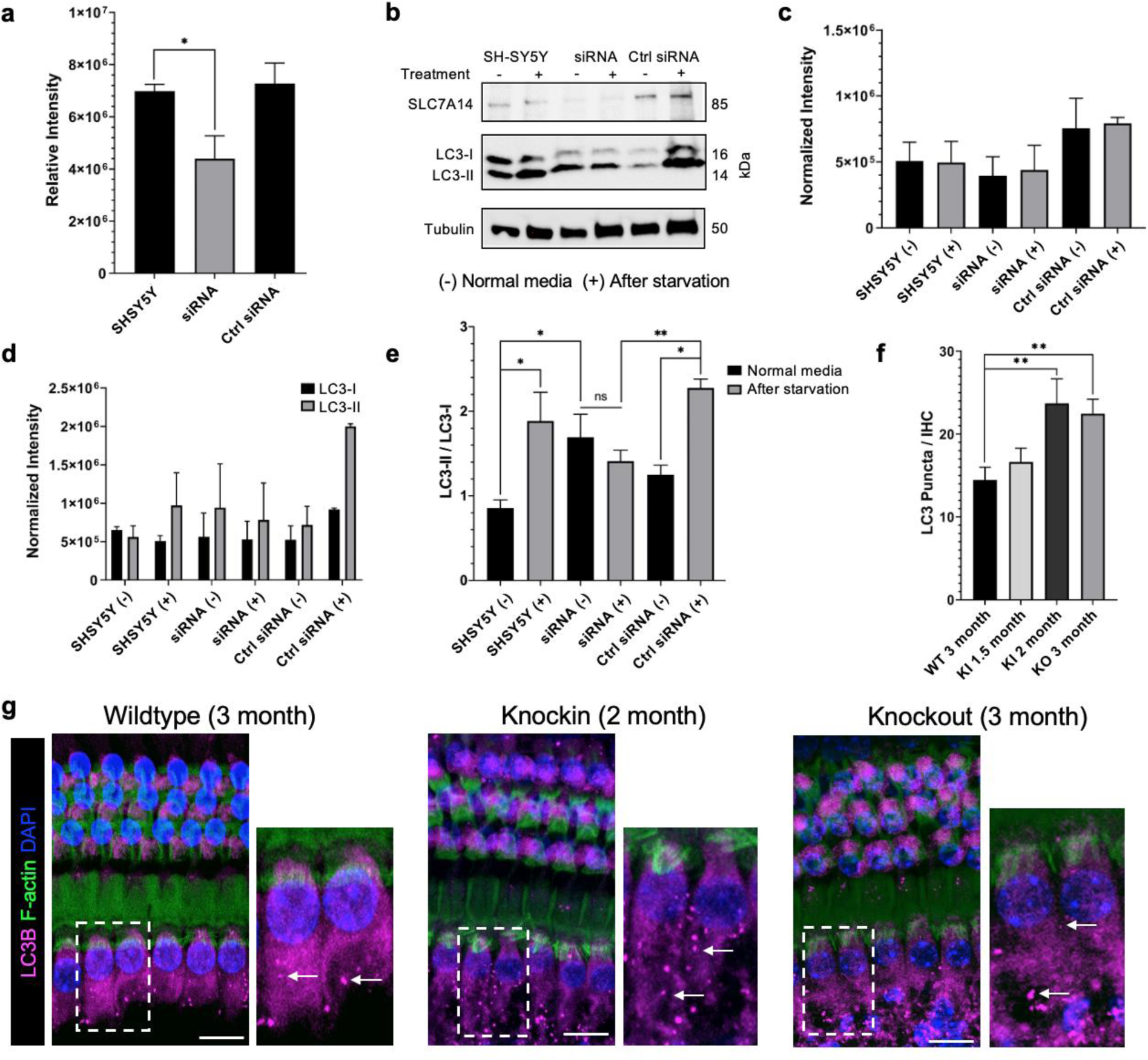
Autophagy activity in SLC7A14 mutants. (a) Quantification of SLC7A14 mRNA expression after gene knockdown in untreated, *SLC7A14* siRNA treated, and control siRNA treated SH-SY5Y cells. Relative band intensity for each sample normalized to GAPDH control band. (b) Western blot image for protein quantification in SH-SY5Y cells treated with siRNA. (c) Quantified SLC7A14 intensity level expression from blot in (b) (normalized to tubulin). (d) LC3-I and LC3-II normalized intensity levels. (e) LC3-II / LC3-I ratio. (a-e: n = 3, unpaired t-test, see methods section for detailed statistical information). (f) Average LC3 puncta in wildtype, knockin, and knockout mouse IHCs quantified from images in (g) (n = 18 IHCs from two cochleae per genotype; unpaired, two-tailed t-test). P values shown as follows: * p < .05, ** p < .005, (Error bars: ±SE). (g) Representative images of mouse cochleae to examine autophagy activity, indicated by LC3 labeled red puncta (white arrows) (Bars: 10 μm).

## DISCUSSION

Previously, *SLC7A14* was identified as a causative gene of autosomal recessive RP, though with no apparent syndromic manifestation (15). In this study, we investigated the function of *Slc7a14* in cochlear hair cells. Unlike other SLC proteins that are ubiquitously expressed in many tissue types, expression of SLC7A14 and corresponding homologs is highly conserved and restricted to distinct neurosensory cell populations including vertebrate inner ear sensory hair cells and retinal photoreceptors. SLC7A14 can serve as a new, highly specific marker for all hair cells in nonmammals and mature IHCs in mammals. *Slc7a14* was detected in developing vestibular hair cells and neonatal OHCs, but no protein expression was observed in adults. This transient expression during development is not unusual as other SLC family members such as *Slc17a8* (vGLUT3), *Slc1a3* (GLAST) and *Slc26a5* (Prestin) are expressed in both IHCs and OHCs early in development but show differential expression in mature IHCs and OHCs (gEAR Database). It should be noted that although high expression of *Slc7a14* was observed in early postnatal spiral ganglion neurons, expression decreased during maturation and no positive staining of SLC7A14 protein was detected after P21, suggesting no functional role of SLC7A14 in these cochlear neurons. Additionally, we confirmed SLC7A14 expression in mature retinal photoreceptors and subcortical regions of the brain, including high-level expression in the hippocampus.

Although SLC7A14 was detected in developing IHCs and OHCs, loss of function apparently did not affect differentiation and development of hair cells; hearing thresholds of 1-month-old KO mice were normal. However, loss of SLC7A14 function caused late-onset hearing loss progressing from mild/moderate loss at 3 months to severe hearing loss by 6 months. Elevated hearing thresholds were observed across all frequencies, consistent with the expression pattern of SLC7A14 in all IHCs with no apical to basal gradient. This auditory phenotype was dissimilar to the pattern of progressive hearing loss observed in mouse models with mutations in other functional hair cells genes, such as *CDH23, KCNQ4* and *Eps8L2,* which often proceeds from high to low frequencies (24–26). DPOAE thresholds of KO mice did not significantly differ from those of wildtype littermates, likely due to the lack of SLC7A14 expression in adult OHCs. Morphological examination of *Slc7a14* KO cochleae with SEM and confocal microscopy confirmed degeneration of IHC stereocilia bundles, decreased MYO7A expression and loss of IHCs, while OHCs remained normal. Thus, loss of *SLC7A14* results in progressive hearing loss due to loss of IHC function. This distinctive type of sensorineural hearing loss, known as auditory neuropathy, is a condition that affects neuronal processing of auditory stimuli due to dysfunction of IHCs, ganglion neurons, or synaptic transmission, while OHC function is sustained (27). Other IHC-specific transporters have also been implicated in pre-synaptic auditory neuropathy including *SLC19A2* (high-affinity thiamine transporter), *SLC17A8* (vesicular glutamate transporter VGLUT3) (28), *OTOF* (synaptic vesicular release) (29, 30), and *CACNA1D* (voltage-gated L-type calcium channel) (31). However, *SLC7A14* is the first intracellular, lysosomal transporter implicated in pre-synaptic, sensorineural hearing loss.

We also examined the auditory function of *Slc7a14* KI mice with the single point mutation SLC7A14 p.Gly330Arg identified in autosomal recessive RP patients. The KI mice exhibited late-onset, progressive hearing loss similar to *Slc7a14* KO mice. In addition, the KI mice displayed progressive thinning of the retina resulting from loss of photoreceptors, a retinopathy indicative of RP that was also observed in the KO mice (15). The pathophysiological evidence from KI mice suggests that the mutation of *SLC7A14* can cause syndromic disease, with progressive hearing and vision impairment. To confirm that *SLC7A14* mutation can cause syndromic disease, we performed audiometric testing in three of the four patients diagnosed with autosomal recessive RP with SLC7A14 p.Gly330Arg mutation (15). They showed mild to moderate bilateral hearing loss, with more severe hearing loss at high frequencies. Additionally, the patients had retinopathy and previous diagnosis of RP. Although the rate of progression and severity of the auditory and visual phenotypes varied among patients, it is conceivable that compounding variables such as genetic variants, sex and environment play a role in pathogenesis. Audiometric testing of additional patients with the same and other variants in SLC7A14 would strengthen our conclusion; however, it should be noted that mutations in this gene are rare and so far have been reported only in east Asia populations(15, 32). The hearing loss due to homozygous SLC7A14 p.Gly330Arg mutation is indicative of a loss of function phenotype as heterozygous mice showed no obvious phenotype, suggesting that one copy of the *SLC7A14* gene appears to be sufficient for function and survival of IHCs. However, it needs to be determined if haploinsufficiency is a risk factor for accelerated age-related hearing and vision loss, or increased vulnerability to noise, intense light and ototoxicity.

A distinct feature shared between hair cells and photoreceptor cells is that they both have ribbon synapses which allow the specialized receptor cells to transduce sensory stimuli into an electrical signal that is graded in response to stimulus intensity (33, 34). A ribbon synapse is characterized by the presence of a large electron-dense presynaptic organelle, the synaptic ribbon, that tethers glutamate-containing synaptic vesicles near the active zone. The cell-type specific expression of the lysosomal-associated protein SLC7A14 may cause one to speculate that it is involved in molecular or structural tuning of ribbon synapses via lysosome-mediated vesicle breakdown or signaling. Our study does not support this speculation for two reasons. First, vestibular hair cells also have ribbon synapses with glutamate as a neurotransmitter; however, SLC7A14 is not expressed in vestibular hair cells and not essential for synaptic transmission in vestibular hair cells. Second and more importantly, immunostaining and confocal microscopy showed no measurable decrease in CtBP2 count in *Slc7a14* KO and KI IHCs, suggesting that loss of function does not affect pre-synaptic ribbon structure or vesicle recycling.

To investigate why loss of function caused IHC degeneration, we examined intracellular trafficking and localization of wildtype and mutant SLC7A14. Using colocalization assays and confocal microscopy, we showed that the SLC7A14 transporter is localized to intracellular membranes including the endoplasmic reticulum, Golgi, and trafficked through direct intracellular routes to the lysosome. The mutant SLC7A14 p.Gly330Arg showed reduced membrane targeting, both in transfected cells and KI IHCs, suggesting that the loss of function variant likely altered protein stability and trafficking. Aberrant protein localization disrupts lysosomal function and may also cause cell stress due to cytosolic accumulation of protein. Lysosomes are important regulator of metabolic homeostasis, and dysfunction of lysosomal proteins can trigger complex cellular cascades, disrupt autophagy, and ultimately lead to cell death (35). Normal localization of SLC7A14 to the lysosome membrane, and its function in the uptake of arginine, provides an important clue about the role of the transporter in maintaining cellular homeostasis in that it may facilitate nutrient-sensing mechanisms. Other lysosomal arginine transporters expressed broadly across tissue types, such as SLC38A9, have been shown to act as arginine sensors regulating autophagy and metabolic homeostasis (36–38). Dysfunction of autophagy in post-mitotic cells, whether through prevention of autophagosome formation, loss of lysosomal function, or other associated mechanisms has been implicated in pathogenesis of neurodegenerative disease (39). We showed that decreased expression of SLC7A14 caused an increase in basal autophagy *in vitro*. Furthermore, the KO and KI IHCs both showed an increase in autophagosome formation compared to age-matched, wildtype IHCs. These results hint at dysregulation of autophagy, resulting from *SLC7A14* mutation, as a pathophysiological mechanism that leads to progressive photoreceptor and IHC loss (40, 41). Our study suggests that SLC7A14 may contribute to nutrient sensing lysosomal function rather than macromolecule degradation and cellular recycling. However, it is unclear how SLC7A14-associated lysosomal dysfunction initiates the pathogenic cascades that cause cell degeneration and death. Disruption of autophagy has been shown to cause loss of photoreceptors (40). The IHC-specific expression of SLC7A14 in the cochlea may underlie differential autophagy mechanisms in IHCs to maintain cellular homeostasis and promote cell survival, compared to the functionally distinct and more vulnerable OHCs. The heterogeneity of lysosomes in IHCs and OHCs, including both the IHC-specific SLC7A14 expression and differential expression of other lysosomal proteins, including LAMP2, suggests both common and distinctive lysosome functions in cochlear hair cells.

Investigation of the pathogenesis of LSDs has elucidated more diverse functions of lysosomes, now considered to be a key cellular signaling organelle regulating processes from metabolic homeostasis to membrane repair, immune response, and cell survival (21). The majority of the over 70 LSDs present as progressive neurological diseases associated with accumulation of a variety of macromolecules due to dysfunction of lysosomal enzymes or transporters (4). While LSDs with a distinctive auditory phenotype are rare, studies from two LSD mouse models with Mucopolysaccharidosis type I or type VII showed progressive hearing loss from 2 months of age (42, 43). However, this hearing loss was not due to degeneration and loss of hair cells but rather alterations in mass and stiffness of the cochlear structures. A recent study showed that mice lacking expression of two lysosomal cation channels, mucolipins 1 and 3, displayed accelerated age-related hearing loss due to OHC degeneration (44). Unlike our observations in SLC7A14 KO hair cells, there was no observed increase in lysosome-mediated autophagy activity (LC3), rather the absence of both mucolipins caused swelling and increased permeabilization of lysosomes which led to cell death. While these studies also underscore the importance of lysosome function in hair cell survival, the current study is the first to show that lysosomal-autophagy dysfunction causes syndromic hearing and vision impairment in mice and humans.

More than 120 genes have been associated with hearing loss with some related to syndromic disease including Usher, Alport, and Stickler syndromes (45, 46). While mutations of *SLC7A14* cause hearing loss and RP, similar to Usher syndrome, there are some differences between them. First, all ten genes (*CDH23, CIB2, CLRN1, DFNB31, GPR98, MY07A, PCHD15, USH1C, USH1G, USH2A*) identified in Usher syndrome are expressed in both IHCs and OHCs, and encode proteins associated with structure and function of the stereocilia bundle (47). In contrast, SLC7A14 is an intracellular transporter localized to the lysosomal membrane solely in IHCs. *SLC7A14* mutation led to degeneration of IHCs, but not OHCs, producing a hearing loss phenotype characteristic of auditory neuropathy. Second, unlike some of the Usher type I and type II mutations which cause profound congenital deafness, mutation of *SLC7A14* does not affect differentiation and development of hair cells and photoreceptor cells. The onset of hearing and vision loss is progressive, similar to Usher type III. However, unlike Usher type I and III which display a vestibular phenotype, *SLC7A14* mutation does not lead to vestibular disorder, likely due to the lack of SLC7A14 expression in mature vestibular hair cells. Finally, we should add that although we did not examine cognitive function in mice and humans, the specific, high-level expression of *SLC7A14*/SLC7A14 in hippocampal neurons may suggest an important functional role in these neurons. Thus, it is reasonable to speculate that mutations of *SLC7A14* may also affect learning and memory, which would also distinguish *SLC7A14* from other genes in syndromic hearing loss.

In summary, we showed that SLC7A14 is a novel marker for vertebrate hair cells and is highly specific to IHCs in mammals. SLC7A14 is expressed in the lysosomal membrane and loss-of-function mutation causes auditory neuropathy and retinitis pigmentosa in mice and humans. As a non-enzymatic component of the lysosome, SLC7A14 is essential for proper regulation of autophagy and mutation disrupts lysosomal homeostasis, resulting in cell degeneration. Degeneration of both IHCs and photoreceptors suggests similarities in a key homeostatic mechanism in these cell types. This is the first study to show that lysosomal-autophagy dysfunction causes syndromic hearing loss in mice and humans. SLC7A14 joins the long list of other SLC transporters linked to human disease (48). Recognition of *SLC7A14* as an additional candidate gene in patients with syndromic vision and hearing loss is important for proper prognosis and treatment. Since the degeneration of IHCs and photoreceptors progresses after birth, this is an ideal syndromic disease model for targeted gene therapy to correct pathogenic variants prior to onset of sensory deficits thereby preventing permanent hearing and vision impairment.

## METHODS

### Animal Models

Wildtype male and female C57BL/6J mice were obtained from Jackson Laboratory and bred for gene and protein expression analyses. Knockout and knockin mice aged one month to a year, both male and female, were used for expression assays and electrophysiology experiments. All mouse experiments were approved by the Creighton IACUC or Wenzhou Medical University and Capital Medical University in China. Other species used in vertebrate expression experiments included *Tg(Pou4f3:GFP*) zebrafish [Creighton University], brown leghorn chicken and red-eared slider turtle [Stanford University], and Sprague Dawley rat [NIH/NIDCD]. All animal procedures were approved by the respective institution animal care and use committees.

The knockout mice were generated previously (15). Heterozygous mice were bred to obtain *Slc7a14*^+/+^ (wildtype), *Slc7a14*^+/-^ (heterozygous), and *Slc7a14*^-/-^ (KO) mice. Genotyping was conducted using genomic DNA extracted from tissue samples. PCR reactions were performed using *Taq* PCR master mix kit (Qiagen) with the following primers: forward: 5’ TCTATCAGCAAACCTTCACTGCAAC-3’ and reverse: 5’ CTGTCAATCATACTGTCAACATGGGTTC-3’.

Because the distinct knockout phenotype is a result of only a 10 base pair deletion, Sanger sequencing of the ∼550 base pair (bp) product followed by genomic alignment confirmed the genotype of experimental mice (Supplementary Fig. 3). Loss of SLC7A14 expression was confirmed by antibody-mediated immunofluorescence.

The knockin (KI) mice were generated using CRISPR-Cas9 mediated genomic recombination. DNA donor sequences, encoding exon 6 with the human mutant codon c.AGG (p.Gly330Arg), together with Cas9 nickase and sgRNA were injected into C57BL/6 mouse zygotes. Full sequences are available upon request. Founder mice (F0) were screened for the substitution and then outcrossed with wildtype mice to generate a stable F1 generation. DNA sequencing was performed to confirm that the target gene had the correct point mutation encoding SLC7A14 p.Gly330Arg (Supplementary Fig. 3). Heterozygous mice (*Slc7a14*^+/c.AGG^) were mated to generate the experimental genotypes including wildtype (*Slc7a14*^+/+^) and homozygous KI mice (*Slc7a14*^c.AGG/c.AGG^). Mice were genotyped with forward (5’-CGTATGTGTCTGTGAGCATGA-3’) and reverse (5’-CAAGGACGGCAGGTTTTTGG-3’) primers and the missense mutation was verified by Sanger sequencing.

### Human subjects

Patient participants with RP were identified and clinically diagnosed in a previous study. For this study, further ophthalmic examination and auditory tests were conducted with informed consent of each participant. The protocol was approved by the ethics committees of Wenzhou Medical University and Capital Medical University.

### Sample processing and histology

Mice were anesthetized with a ketamine/xylaxine mixture then decapitated. Tissues of interest, including the brain, eyes, and otic capsules were removed and placed in 4% paraformaldehyde (PFA) in 1X phosphate buffered saline (PBS) for 24 hours at 4 °C. Adult cochlea were decalcified in 120 mM EDTA for 3 to 5 days at 4 °C until the bony tissue was soft and flexible. Fixed tissues were washed in 1X PBS three times, then dehydrated using a standard ethanol series, embedded in paraffin, and sectioned (8 - 15 µm thick). The brain, retina, and cochlear sections were used for subsequent gene and protein expression assays.

### RNAscope

The RNAscope® based small molecule fluorescent in situ hybridization (smFISH) assay from Advanced Cell Diagnostics (ACD) was used to examine *Slc7a14* mRNA expression in the developing mouse cochlea. Samples were pretreated according to the ACD protocol for formalin-fixed paraffin embedded tissue. However, due to the fragility of the tissues, the pretreatment target retrieval and protease steps were modified. Tissues were covered with freshly prepared 0.5% pepsin + 5 mM HCl in deionized water and incubated in the humidifying chamber at 37 °C for 10 min. The remaining protocol was conducted according to the RNAscope 2.5 HD Detection Reagent RED user manual and published protocols (49). Following the final wash, samples were immunolabelled using the protocol described below to label hair cells using anti-MYO7A primary antibodies. Probes were detected by red channel fluorescence, with no probe treatment included as a negative control.

### Immunofluorescence

Mouse tissues prepared as described above were used for immunofluorescence experiments. Formalin-fixed paraffin embedded tissues were sectioned then deparaffinized and rehydrated. Additional otic capsules were dissected for whole mount preparations. Samples were washed in 1X PBS, then permeabilized and blocked in 1X PBS with 0.2% Triton X-100 (PBS-T) and 5% normal goat serum (NGS), followed by incubation with primary antibodies (Table 1) overnight at 4 °C. Samples were rinsed in 1X PBS three times and then incubated with Alexa-Fluor (AF) conjugated secondary antibodies (Table 1) for one hour at room temperature followed by subsequent washes. For some samples, additional staining with 4′,6-diamidino-2-phenylindole (DAPI) or AF488-Phalloidin (anti-F-Actin) was used to label nuclei and stereocilia, respectively. Finally, samples were rinsed three times in 1X PBS and mounted with SlowFade (Invitrogen), then imaged.

**Table 1:** Antibodies

| Antibody Target | Supplier | Catalog Number | Working Concentration |
| --- | --- | --- | --- |
| **Primary Antibodies** |  |  |  |
| Anti-SLC7A14 | Sigma | HPA045929 (lot:R43519) | 1:400 (IF);<br>1:1000 (WB) |
| Anti-GLG1 | R & D Systems | AF7879 | 2.0 µg/mL |
| Anti-LAMP1- mouse | DSHB | 1D4B | 2.0 – 5.0 µg/mL |
| Anti-LAMP1- human | DSHB | H4A3 | 2.0 – 5.0 µg/mL (IF); 0.5 µg/mL (WB) |
| Anti-LAMP2- mouse | DSHB | GL2A7 | 2.0 – 5.0 µg/mL |
| Anti-LAMP2- human | DSHB | H4B4 | 2.0 – 5.0 µg/mL |
| Anti-Calreticulin (D3E6) XP Rabbit mAb Alexa Fluor 594 | Cell Signaling Technology | 77344 (lot:1) | 1:100 |
| Anti-LC3B (D11) XP Rabbit mAb | Cell Signaling Technology | 3868 (lot:13) | 1:200 (IF);<br>1:1000 (WB) |
| Anti-MYO7A | Proteus Biosciences | 25-6790 (lot:110119) | 1:1000 |
| Anti-α-Tubulin | Abcam | ab4074 | 1:400 |
| Anti-CtBP2 | BD Transduction Laboratories | Lot: 612044 | 1:300 |
| Anti-Sox2 | Santa Cruz Biotechnology | sc-365823 | 1:100 |
| Anti-Neurofilament | DSHB | 2H3 | 2.0 – 5.0 µg/mL |
| **Secondary Antibodies** |  |  |  |
| Alpaca anti-rabbit IgG recombinant VHH, AF488 | Chromotek | srbAF488-1 (lot:90221043AF1) | 1:400 |
| Alpaca anti-rabbit IgG recombinant VHH, AF568 | Chromotek | srbAF568-1 (lot:90222043AF3) | 1:400 |
| DAPI | Sigma | D9542 (lot:127M4055V) | 1:10000 |
| Goat anti-mouse IgG, HRP | Abcam | Ab205719 (lot:GR3279214-1) | 1:10000 |
| Goat anti-rabbit IgG, HRP | Abcam | Ab205718 (lot:GR3269880-6) | 1:10000 |
| Goat anti-mouse, AF488 | Invitrogen | A-32723 | 1:1000 |
| Goat anti-mouse, AF568 | Invitrogen | A11004 (lot:1906485) | 1:1000 |
| Goat anti-rabbit, AF568 | Invitrogen | A11077 (lot:2110844) | 1:1000 |
| Goat anti-rabbit, AF633 | Invitrogen | A21070 (lot:2079350) | 1:1000 |
| Donkey anti-sheep, AF633 | Invitrogen | A21100 (lot:1871971) | 1:1000 |

Tissues from vertebrate species were collected following euthanasia according to respective institutional animal care and use protocols. Zebrafish *Tg(Pou4f3:,GFP*) larvae (72 hpf) were euthanized and fixed in 4% PFA in PBS overnight at 4 °C. Immunostaining was conducted as previously described (50). Red-eared sliders were decapitated, the lower jaw removed, head bisected, brain removed, and bone of the ear capsule dissected away to allow penetration of fixative. The bisected heads were stored in 4% paraformaldehyde in PBS at room temperature overnight. The papilla was then dissected out of the bony capsule in PBS. Fertilized chicken eggs were purchased from AA Laboratory Eggs Inc. (Westminster, CA). Following euthanasia and decapitation, post hatch day 7 chicken heads were bisected, brain and lower jaw removed, and the bony ear capsule (containing all auditory and vestibular organs) was excised with surgical scissors. Following dissection to expose the basal region the tissue was placed in 4% PFA at room temperature for 1 hour. Immunostaining of chicken and turtle tissues was conducted as previously described (51), modifying the protocol to accommodate free-floating vibratome sections, which were stained in mesh-bottom baskets (52). Rat cochleae were prepared similarly to the mouse cochlea as described above. Human cochlear sections were kindly provided by the NIDCD National Temporal bone laboratory at UCLA. Celloidin embedded human inner ear sections were immunostained using a published protocol (53). DAPI was used to label the nuclei of inner ear cells and anti-SLC7A14 detected cell-specific protein expression.

### Confocal microscopy

Visualization of smFISH and immunofluorescence was conducted using Zeiss laser scanning confocal microscopes LSM700, 710 or 880. Sections were imaged at 1.0 zoom at 40x or 63x magnification using Zen Black acquisition software. Additionally, samples for colocalization analyses, included cultured cells, were imaged on a Nikon Ti-E with a Yokogawa Spinning Disc Confocal with a Flash 4.0 Hamamatsu Monochrome camera.

### Scanning electron microscopy

The protocol described by Jia et al. (2009) was referenced for sample preparation and imaging protocol (54). Briefly, cochleae from wildtype and knockout mice were fixed in 2.5% glutaraldehyde in 0.1 M sodium cacodylate buffer (pH 7.4) containing 2 mM CaCl2 for 24 hours at 4 °C. The cochleae were post-fixed for one hour in 1% OsO4 in 0.1 M sodium cacodylate buffer. Samples were then dehydrated in ethanol, critical point dried from CO2 and sputter-coated with gold. SEM was conducted on a FEI Quanta 200 at the University of Nebraska Medical Center imaging core and images were acquired digitally.

### Auditory electrophysiology and retinography

To characterize the auditory phenotype of the mice we measured both the auditory brainstem response (ABR) and distortion product otoacoustic emission (DPOAE) using our established protocols (7, 55). ABRs measurements were elicited with standard tone bursts from 4 to 50 kHz. The response signals were amplified (100,000x), filtered and acquired by a TDT RZ6 (Tucker-Davis technologies). Each averaged response was based on 500 stimulus repetitions. DPOAE thresholds were elicited by input of two primary tones of different frequencies (*f*1 and *f*2) from two electrostatic speakers, with the f2 level 10 dB lower than the f1 level (Tucker-Davis Technologies EC1 phones). The sound pressure obtained from the microphone in the ear-canal was amplified and Fast-Fourier transforms were computed from averaged waveforms of ear-canal sound pressure. The DPOAE response was measured in response to *f*1 and *f*2, with f2/f1 = 1.2. Recorded ABR and DPOAE thresholds were graphed, and statistical analysis was conducted using GraphPad Prism.

To test the auditory function in diagnosed autosomal recessive RP patients, otoscopy and tympanometry were performed to examine the ear canal and middle ear, and pure tone audiometry was conducted according to standard protocol (56). Normal air conduction threshold search screening was conducted by presenting pure tones from 250 Hz to 8 kHz at the upper limits of human hearing. Decreasing intensities, at 10 dB intervals, were presented until the patient no longer responded. If no deficit was detected up to 8 kHz, extended range testing up to 16 kHz was conducted. Hearing thresholds were recorded for both the left and right ears and compared to age-matched average hearing thresholds (57). DPOAE measurements of both left and right ears from human patients were conducted based on similar principles described above. The DP-gram was generated with standard clinical level separation with F1 level set to 65 dB and F2 at 55 dB, with the frequency ratio of 1.2. The signal-to-noise-ratio was calculated as mean noise floor with response variability (standard deviations). Details for ophthalmic examinations, including fundoscopy, OCT and EGR, were described in previous publication (15).

### *In vitro* experiments Cell culture

The SH-SY5Y human bone marrow neuroblast cell line (ATCC^®^ CRL-2266™) was selected for *in vitro* assays based on endogenous expression of *SLC7A14*. Cells were cultured according to the product data sheet from ATCC and grown in DMEM: F12 medium supplemented with 10% FBS. Cells were maintained in a humidified 37 °C incubator with 5% CO2 and the culture medium was refreshed every 3 to 4 days. Adherent, differentiated cells resembling neurons were used for all experiments described below.

### Cell transfection

The pcDNA3.1+C-eGFP plasmids (Genscript) containing the wildtype *SLC7A14* (WT pcDNA3.1) or mutant *SLC7A14* c.988G>A (MT pcDNA3.1) were amplified using the high efficiency transformation protocol (NEB). Transformed cells were selected by ampicillin resistance, and amplified in liquid LB + ampicillin media, then incubated at 37 °C for 18 hours in a shaker. Plasmids were isolated using the QIAprep Spin Miniprep Kit (Qiagen) and concentration was measured by Nanodrop. Plasmids were sequenced by Sanger sequencing using the mammalian CMV promoter and aligned to the reference sequence to confirm the correct ORF sequences for both the wildtype and mutant plasmids.

For transfection experiments, SH-SY5Y cells were seeded into 6-well plates and incubated until they were 75% confluent. One hour prior to transfection, the cell media was replaced with Opti-MEM media. Cells were transfected using Lipofectamine3000 (Thermo Fisher) reagent with approximately 2.0 ug/µl of WT or MT plasmid per well. The transfection media was added to the well plates and incubated for 6 hours. After the incubation period, cells were supplemented with normal media and incubated under normal conditions for 48 hours, then screened for transfection efficiency.

### Immunofluorescence

For *in vitro* immunofluorescence experiments, SH-SY5Y cells were seeded at 0.5 x 10^6^ in glass bottomed 35 mm dishes and incubated overnight in normal culture conditions. Media was replaced with 4% PFA and cells were incubated for 2 hours at 4 °C. After fixation the cells were rinsed three times with 1X PBS and then incubated with 0.2% TritonX-100 + 5% NGS for 20 minutes. Primary antibodies targeting proteins of interest (Table 1) were incubated overnight at 4 °C. Following repeat 1X PBS washes, samples were incubated with conjugated secondary antibodies (Table 1) for 2 hours at room temperature. Cells were incubated with DAPI for 10 minutes, washed, and then covered with a 50% glycerol/ 50% 1X PBS solution, then stored at 4 °C prior to imaging. Cells were imaged on a Nikon Ti-E with a Yokogawa Spinning Disc Confocal with a Flash 4.0 Hamamatsu Monochrome camera.

### Colocalization analysis

Live cells were examined for localization of SLC7A14-eGFP to the lysosome. Following transfection, live cells were labelled with LysoTracker® Red DND-99 (Thermo Fisher) at a working concentration of 75 nM. The red fluorophore is acidotropic, which labels the acidic lumen of the lysosomes. Addition of Hoechst 33342 (5 µM) stained the nuclei of the live cells. Transfected cells were incubated with the labelling mixture for one hour at 37 °C, washed with 1X PBS, then supplemented with normal media.

Colocalization assays were conducted both *in vitro* in cultured SH-SY5Y cells and *in vivo* using cochleae from P30 wildtype C57BL/6 mice. Additionally, cochleae from the knockin mice were used to examine localization of the mutant SLC7A14 p.Gly330Arg protein. Protocols for immunofluorescence of these cells and tissues are described above. It is important to note that different LAMP1 and LAMP2 antibodies were used to target human and mouse proteins in SH- SY5Y cells and cochlear tissue. Additionally, the Calreticulin primary antibody was conjugated therefore no secondary antibody incubation was required.

Samples were imaged on a Nikon Ti-E confocal microscope and the colocalization function in the NIS Element software (Nikon) was used to quantify the colocalization of the green (SLC7A14) and red (protein label of target organelle). Thresholding of each fluorophore, as well as selecting a region of interest in the image, allowed for subtraction of background signal to maximize the signal for each channel. Pearson’s correlation coefficient (PCC), a statistical measure of the co-occurrence of the respective fluorophores in each pixel, was used to determine colocalization (18). The PCC measure for each layer in the confocal z-stack image was calculated separately, exported to Excel, and then averaged for each sample. As a result, each co-labeled sample (n = 3 per genotype, per target) was imaged in at least to 3 locations and each image consisted of 10 – 15 layers. Therefore, the average represented at least 100 calculated PCC values.

### Gene knockdown

*SLC7A14* gene knockdown was achieved using siRNA treatment. One day prior to treatment SH-SY5Y cells were seeded in 24-well plates at 0.05 x 10^6^ cells. The target *SLC7A14* siRNA or negative-control siRNA was diluted in the Accell delivery media (Dharmacon). Normal media was replaced with 500 μl of the siRNA mixture for a final concentration of 1 µM per well. Following incubation for 72 hours, the cells were collected, and total RNA or protein was isolated and analyzed as described below.

### RT-PCR

Total RNA from cultured cells was isolated using the RNeasy mini kit (Qiagen) and concentrations were measured using a Nanodrop 2000. Replicate cDNA libraries were constructed from 1 µg of total RNA using the QuantiTect reverse transcription kit (Qiagen). RT-PCR was performed to detect expression of *SLC7A14* (forward primer 5’- CACGGCACATGGAACTAAGC-3’, reverse primer 5’-TGATTCTTCACCTTTCCCCAGG-3’) and *GAPDH* (forward primer 5’-AAGACGGGCGGAGAGAAACC-3’, reverse primer 5’-GGGGCAGAGATGATGACCCT-3’). The reaction proceeded as follows: 94 °C for 4 minutes; 94 °C for 30 seconds, 62 °C for 30 seconds, and 72 °C for 1 minute x 35 cycles; and, 72 °C for 10 minutes. The PCR products were separated by electrophoresis on a 2% agarose gel with SYBRsafe DNA gel stain (Invitrogen), followed by UV imaging to detect PCR products. Band intensities were quantified using Bio-Rad Image Lab 6.1 software and normalized to GAPDH.

### Western blot

Protein expression in cultured cells was quantified by Western blot. Samples were washed with cold PBS and then isolated in RIPA cell lysis buffer and 1X Halt protease inhibitor cocktail (Thermo Fisher). Protein concentration (µg/ml) was measured using the Pierce rapid gold BCA protein assay kit (ThermoFisher) and a Varioskan LUX plate reader (3020-80010) to quantify absorbance at 480 nm. Samples were run on a mini-PROTEAN TGX-stain free gel (Bio Rad) with a 4 to 20% gradient to separate proteins, along with molecular weight markers. The gel was run at 200 V for 30 minutes then imaged. Transfer was completed using the Trans blot turbo system (Bio-Rad), followed by activation and imaging to ensure transfer. The membrane was washed in TBS-T (1X tris buffered saline + 1 % Tween20) and then blocked in EveryBlot blocking buffer (Bio-Rad). The membrane was incubated with primary antibodies (Table 1) in blocking buffer overnight at 4 °C. Following four washes in TBS-T, the blot was incubated with HRP-conjugated secondary antibodies (Table 1) for one hour at room temperature. Following additional wash steps, the membrane was incubated with the Chemiluminescence substrate (Bio-Rad) for 5 minutes, then imaged. Quantification of the bands of interest was conducted using Bio-Rad Image Lab 6.1 software.

### Autophagy activity

After confirming successful gene knockdown, we repeated the experiment to measure changes in autophagy activity after decrease of SLC7A14 expression. The starvation treatment was simply a two-hour incubation in Hank’s buffered saline solution (HBSS) which successfully increased autophagy in wildtype SH-SY5Y cells. Immediately following the 72-hour incubation in the knockdown media, the cells were supplemented with normal media for 2 hours to ensure that autophagy activity was not upregulated due to the reduced serum Accell media used for knockdown. The cells were then washed twice with PBS, followed by incubation in HBSS for 2 hours. Cells were harvested for total RNA or total protein (described above), and autophagy activity was analyzed by Western blot (see protocol above; antibodies listed in Table 1). The expression of proteins of interest, SLC7A14 and LC3B, were normalized to α-tubulin intensity. Autophagy activity was measured based on the LC3-II to LC3-I expression ratio, which indicates active formation of autolysosomes.

Autophagy *in vivo* was examined using immunofluorescence of the LC3B protein(58). To specifically examine basal autophagy, mice were not subjected to noise exposure (ie. ABR testing) or stress-inducing starvation immediately prior to tissue harvesting. The wildtype, knockout, and knockin mice cochleae (aged 1.5 to 3 months) were immunostained with anti-LC3B antibody, phalloidin-AF488 and DAPI, then imaged with a Zeiss 710 confocal. The number of puncta per IHC was quantified in 3 locations per cochlea using Image J.

### Statistical analysis

All statistical analysis was conducted in GraphPad Prism. The auditory physiology measurements were analyzed by two-way ANOVA with repeated measures, grouped to include biological replicates, followed by a post-hoc analysis Tukey’s multiple comparisons test (α = .05).

Knockout mouse analysis (n = 6, all ages): ABR: F = 223, df = 2, p < 0.0001; ABR shift: F = 32.4, df = 2, p < 0.001; and, DPOAE: F = .821, df = 2, p = 0.4431. Knockin mouse analysis (2-month n = 6, 3.5-month n = 3): 2 month ABR: F = 9.057, df = 2, p = 0.0002; 3.5 month ABR: F = 116.9, df = 2, p < 0.0001; and, 3.5 month DPOAE: F = 0.00, df = 2, p > 0.9999. P values from post-hoc analysis presented in figure are adjusted p values. Mean PCC values from *in vitro* colocalization analysis were analyzed by one-way ANOVA, F = 61.72, df = 3, p < 0.0001. Post-hoc analysis was conducted with Tukey’s multiple comparisons test (α = .05) and significance reported as adjusted p values. Colocalization studies of wildtype and knockin mouse cochleae samples (n = 3 per genotype) were analyzed by unpaired T-test: Calreticulin: t = 23.32, df = 339, p < 0.000001; LAMP1: t = 16.97, df = 181, p < 0.0001; LAMP2: t = 7.671, df = 194, p < 0.0001. Following gene knockdown in transfected cells the differences in mRNA expression, quantified as normalized intensity, were analyzed by unpaired T-test (n = 3, df = 4): Control vs. *SLC7A14* siRNA: t = 2.81, p = 0.048; Control vs. negative Control siRNA: t = 0.349, p = .744; Control vs Accell media alone: t = 0.647, p = 0.553. Protein expression quantified by western blot was compared by one-way ANOVA (n = 2) and no significance was found among the experimental groups (F = 1.089, p = 0.4516). Autophagy activity (LC3II: LC3I ratios) *in vitro* were analyzed by unpaired, two-tailed t-test (n = 3, df = 4): SH-SY5Y control fed vs. starved: t= 2.899, p = 0.044; siRNA fed vs. starved: t = 0.9328, p = 0.4037; Control siRNA fed vs. starved: t = 6.627, p = 0.0027; siRNA fed vs. SH-SY5Y control fed: t= 2.899, p = 0.044; SH-SY5Y control fed vs. Control siRNA fed: t = 2.669, p = 0.0559. Quantified LC3 puncta in mouse IHCs were also compared with unpaired, two-tailed t-test (n = 18 IHCs per genotype, df = 32): Wildtype vs. Knockin (1.5 month): t = 0.9648, p = 0.3419; Wildtype vs. Knockin (2 month): t = 2.765, p = 0.0094; Wildtype vs. Knockout (3 month): t = 3.460, p = 0.0016.

## Data Availability

All data generated in this study are included in the publication. Hearing and vision exams data from patients will be available upon request when the manuscript is accepted.

## Acknowledgements

This work was supported by NIH grant R01 DC016807 and the Béllucci Depaoli Family Foundation to DH. ZBJ is partly supported by the Beijing Natural Science Foundation (Z20J00122) and National Natural Science Foundation of China (81970838). YL is supported by National Natural Science Foundation of China (#81870718 and #81770996). We would like to thank Dr. Holly Stessman for the kind gift of the SH-SY5Y cells. We also want to thank Dr. Ivan Rebustini at NIH-NIDCD for imaging rat tissues and Mike Schnee at Stanford University for isolating turtle papilla. The Imaging and Molecular Biology Cores at the Translational Hearing Research Center of Creighton University School of Medicine are supported by grant NIH grant 1P20GM139762-01 from the NIGMS and by Béllucci Depaoli Family Foundation. Human cochlear samples were provided by the NIDCD National Temporal bone laboratory at UCLA supported by NIDCD/NIH Grant U24: 1U24DC015910-01.

## Author contributions

KG, ZBJ and DH designed experiments. KG, HZL, YL, XCZ, AJ, SSG, YL, TW, BK and DH performed auditory function tests, immunostaining, imaging, and in vitro molecular biology experiments in cell lines. ZBJ and YL provided knockout and knockin mouse lines. BPC and DH tested human patient hearing function. ZBJ, RJS and CJZ tested vision in patients. HZL, TW, CJZ and DH examined mouse retina morphology. KG and DH wrote the manuscript.

