## Supplemental Figures for "Mutation of SLC7A14 Causes Auditory Neuropathy and Retinitis Pigmentosa Mediated by Lysosomal Dysfunction"

**Supplemental Material for Giffen et al on SLC7A14**

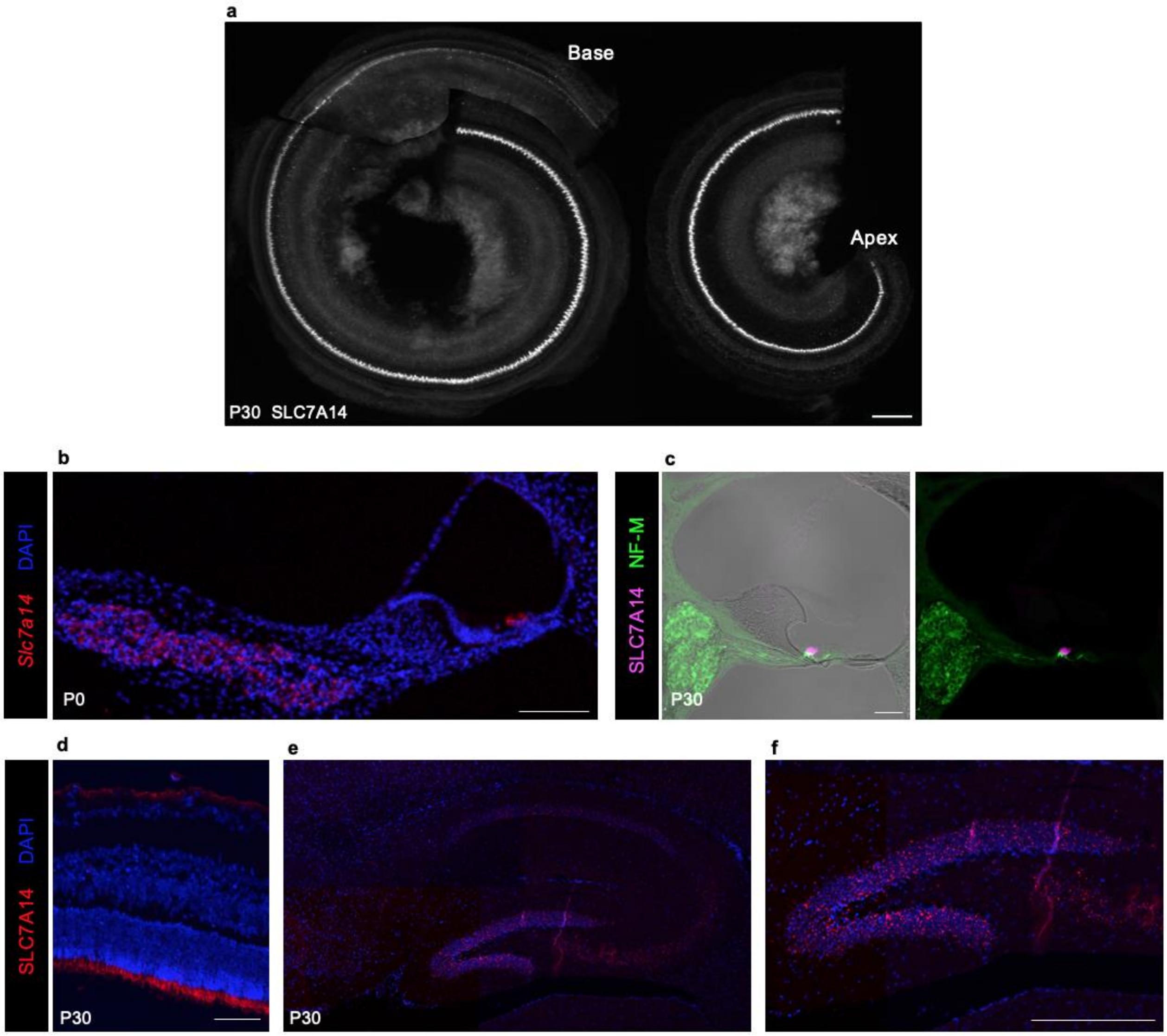


**Supplementary Fig. 1**: SLC7A14 expression in mouse tissues. (a) Expression of SLC7A14 in IHCs extending from the base to the apex of the cochlea (Bar: 100 µm). (b) *Slc7a14* expression detected by RNAscope in early postnatal mouse organ of Corti and spiral ganglion (Bar: 100 µm). (c) SLC7A14 protein expression was not observed in the spiral ganglion neurons (Neurofilament positive) at any age (P30 shown above) (Bar: 50 µm). (d) Confirmation of SLC7A14 expression in the photoreceptor layer of the adult mouse retina and retinal ganglion neurons (Bar: 50 µm). (e) Expression of SLC7A14 in the mouse hippocampus, and (f) highly specific expression in the dentate gyrus (Bar: 500 µm).

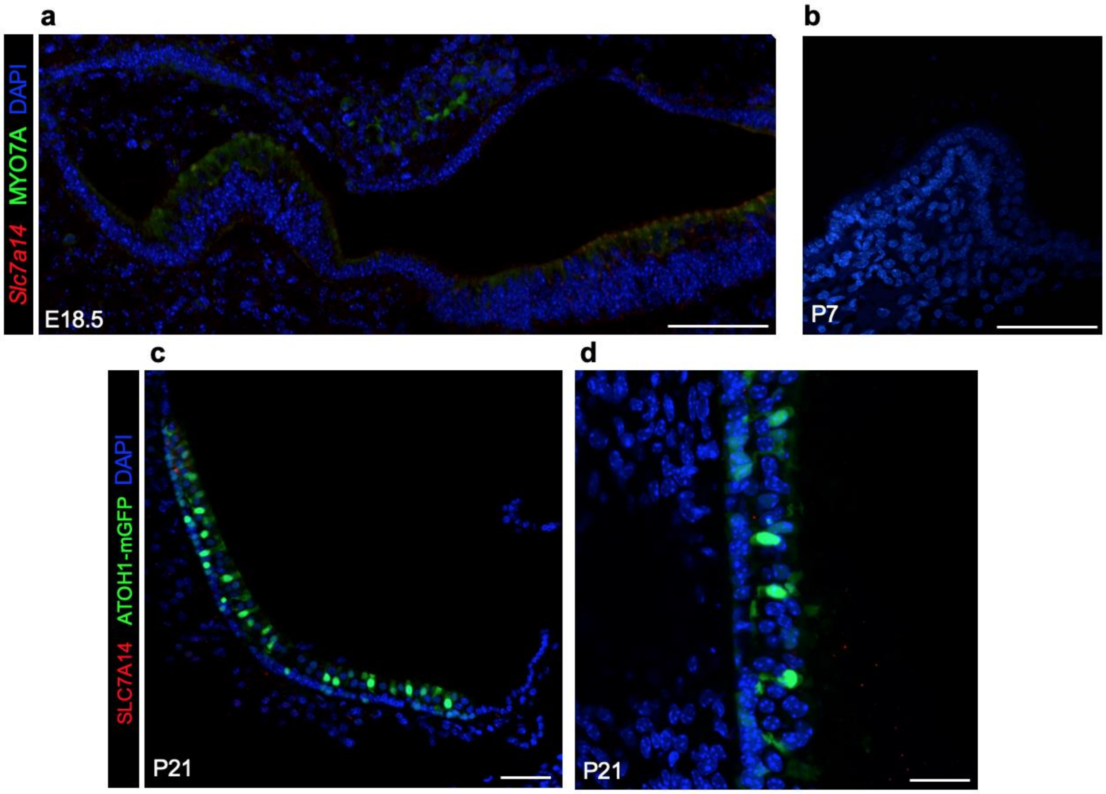


**Supplementary Fig. 2**: Expression of *Slc7a14*/SLC7A14 in developing vestibular organs. RNAscope smFISH of *Slc7a14* (red probe) in developing mouse vestibular hair cells at (a) embryonic day 18.5 and (b) postnatal day 7 (Bars: 50 µm). Adult mouse (Atoh1:mGFP) utricle labelled with SLC7A14 antibody. Bars: (c) 50 µm and (d) 20 µm.

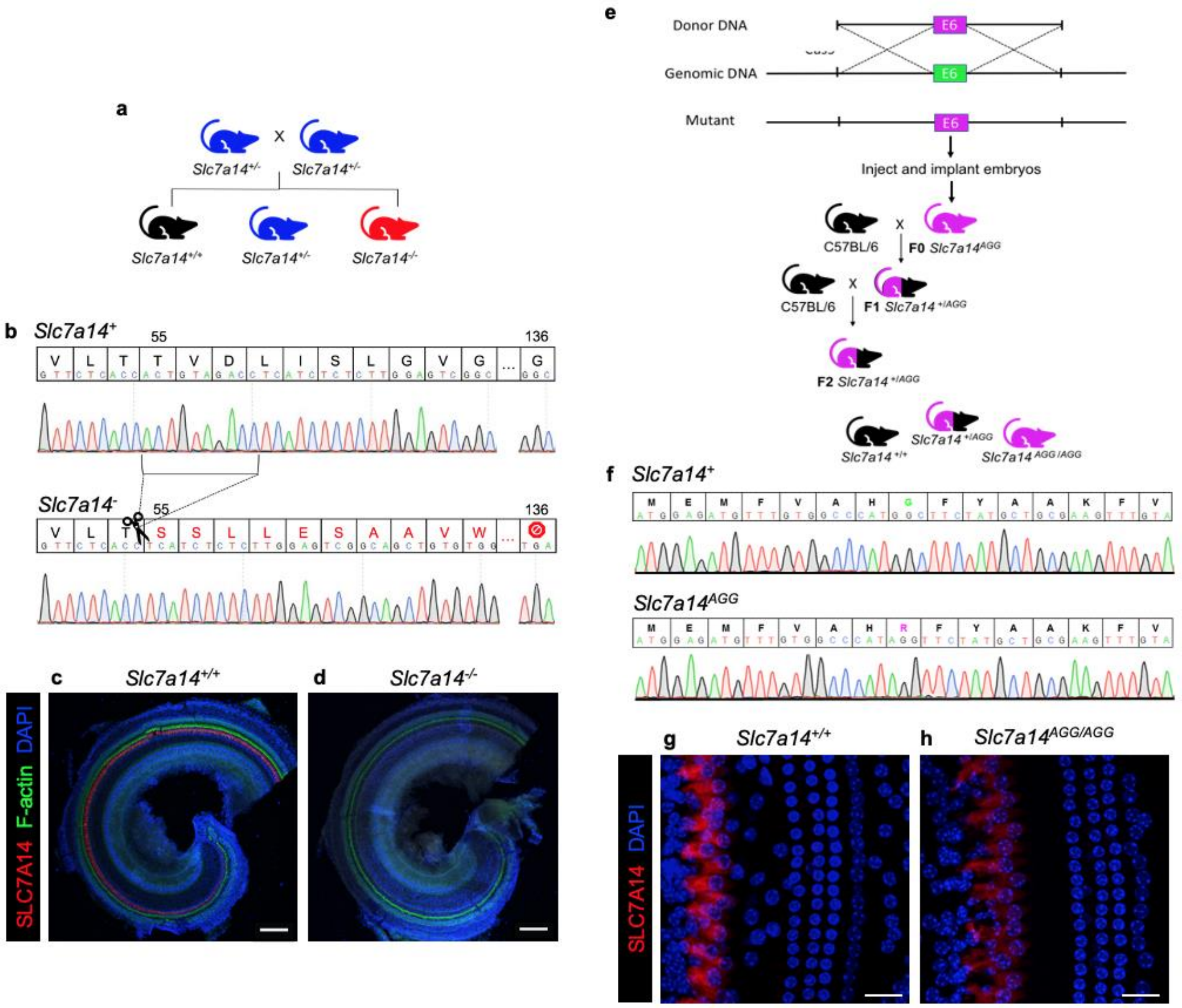


**Supplementary Fig. 3**: Transgenic mouse lines. (a) Breeding scheme for generating knockout mice. Heterozygous mice were bred to obtain all experimental animals. (b) Genomic DNA was isolated and amplified by PCR. Sanger sequencing of products confirmed the 10 bp deletion in the knockout mouse, compared to the wildtype mouse coding sequence, and the subsequent frameshift and early truncating stop codon at residue 136. Antibody-mediated immunofluorescence confirmed normal SLC7A14 expression (red) in (c) wildtype IHCs, with no visible expression observed in the (d) knockout mouse cochlea (Bars: 100 $\mu$m). (e) The *Slc7a14* knockin mouse line was generated using CRISPR/Cas9 nickase to induce single stranded breaks followed by homology driven repair with a donor template containing exon 6 with the point mutation encoding p.Gly330Arg. Mice were outcrossed to C57BL/6 mice to obtain a F2 generation with the heritable mutation. (f) PCR and Sanger sequencing of products confirmed mice with wildtype or knockin sequences. SLC7A14 expression in (g) wildtype and (h) knockin mouse cochleae (Bars: 20 $\mu$m).
